# Spatiotemporal analysis of surveillance data enables climate-based forecasting of Lassa fever

**DOI:** 10.1101/2020.11.16.20232322

**Authors:** David W. Redding, Rory Gibb, Chioma C. Dan-Nwafor, Elsie A. Ilori, Yashe Rimamdeyati Usman, Oladele H. Saliu, Amedu O. Michael, Iniobong Akanimo, Oladipupo B. Ipadeola, Lauren Enright, Christl A. Donnelly, Ibrahim Abubakar, Kate E. Jones, Chikwe Ihekweazu

**Author notes:** These authors contributed equally to this work (David W. Redding, Rory Gibb).

## Abstract

Lassa fever (LF) is an acute rodent-borne viral haemorrhagic fever that is a longstanding public health concern in West Africa and increasingly a global health priority. Recent molecular studies^1,2^ have confirmed the fundamental role of the rodent host (*Mastomys natalensis*) in driving human infections, but LF control and prevention efforts remain hampered by a limited baseline understanding of the disease’s true incidence, geographical distribution and underlying drivers^3^. Here, through analysing 8 years of weekly case reports (2012-2019) from 774 local government authorities (LGAs) across Nigeria, we identify the socioecological correlates of LF incidence that together drive predictable, seasonal surges in cases. At the LGA-level, the spatial endemic area of LF is dictated by a combination of rainfall, poverty, agriculture, urbanisation and housing effects, although LF’s patchy distribution is also strongly impacted by reporting effort, suggesting that many infections are still going undetected. We show that spatial patterns of LF incidence within the endemic area, are principally dictated by housing quality, with poor-quality housing areas seeing more cases than expected. When examining the seasonal and inter-annual variation in incidence within known LF hotspots, climate dynamics and reporting effort together explain observed trends effectively (with 98% of observations falling within the 95% predictive interval), including the sharp uptick in 2018-19. Our models show the potential for forecasting LF incidence surges 1-2 months in advance, and provide a framework for developing an early-warning system for public health planning.

---

In 2018 and 2019, Nigeria recorded its highest annual incidence of Lassa fever (633 confirmed cases in 2018 and 810 in 2019, across 28 states), prompting national and international healthcare mobilisation and raising concerns of an ongoing, systematic emergence of LF nationally^1,2^. Lassa virus (LASV; Arenaviridae, Order: Bunyavirales) is a WHO-listed priority pathogen and a major focus of international vaccine development funding^3^ and, although often framed as a global health threat, LF is foremost a neglected endemic zoonosis. A significant majority of cases – including those from recent years in Nigeria^4^ – are thought to arise directly from spillover from the LASV reservoir host, the widespread synanthropic rodent *Mastomys natalensis*, although with hospital-acquired infections potentially occurring in small clusters of human-to-human cases^5–7^. Risk factors for spillover, while not well understood, are thought to include factors that increase direct and indirect contact between rodents and people, including ineffective food storage, housing quality, and certain agricultural practices such as crop processing^8,9^. Evidence of correspondence between human case surges and seasonal rainfall patterns suggests that LF is a climate-sensitive disease^10^, whose incidence may be increasing with regional climatic change^11^. The present-day incidence and burden, however, remain poorly defined, because LASV surveillance has historically been opportunistic or focused on known endemic districts with existing diagnostic capacity^12^, and often-cited annual case estimates (of up to 300,000) are consequently based on only limited serological evidence from a handful of early studies^13,14^. This, alongside LF’s nonspecific presentation, means that many mild or subclinical infections (possibly around 80% of cases) are thought to go undetected^15,16^. The patchy understanding of LF’s true annual incidence and drivers hinders disease control^17^ and provides limited contextual understanding of whether the recent surges in reported cases result from improvements in surveillance or a true emergence trend.

To address this, we analyse the first long-term spatiotemporal epidemiological dataset of acute human Lassa fever case data, systematically collected over 8 years of surveillance in Nigeria. This dataset, collated by the Nigeria Centre for Disease Control (NCDC), consists of weekly epidemiological reports of acute human LF cases collected by all 774 local government authorities (LGAs) across Nigeria between January 2012 and December 2019 (Figure 1). Throughout the study period, 161 LGAs from 32 of 35 states reported cases, with a mean annual total of 276 (range 25 to 816) confirmed cases, though with evidence of pronounced spatial and temporal clustering. For example, most cases (75%) are reported from just 3 of the 36 Nigerian states (Edo, Ondo and Ebonyi), with lower incidence overall in northern endemic states, in areas notably distant from diagnostic centres. There is consistent evidence of seasonality in all areas across the reporting period, with the exception of 2014-15, when a lull in recorded cases was coincident in timing with the West African Ebola epidemic (Extended Data Figure 1). Annual dry season peaks of LF cases typically occur in January, confirming past hospital admissions data from Nigeria^18,19^ and Sierra Leone^20^, with some secondary peaks evident in early March and, increasingly, a small number of cases detected throughout the year (Figure 1). Both overall temporal trends and cumulative case curves suggest that 2018 and 2019 appear to be markedly different from previous years, with very high peaks in confirmed cases extending from January into March, and high suspected case reporting continuing throughout 2019 (Figure 1, Extended Data Figure 1).

**Figure 1:**
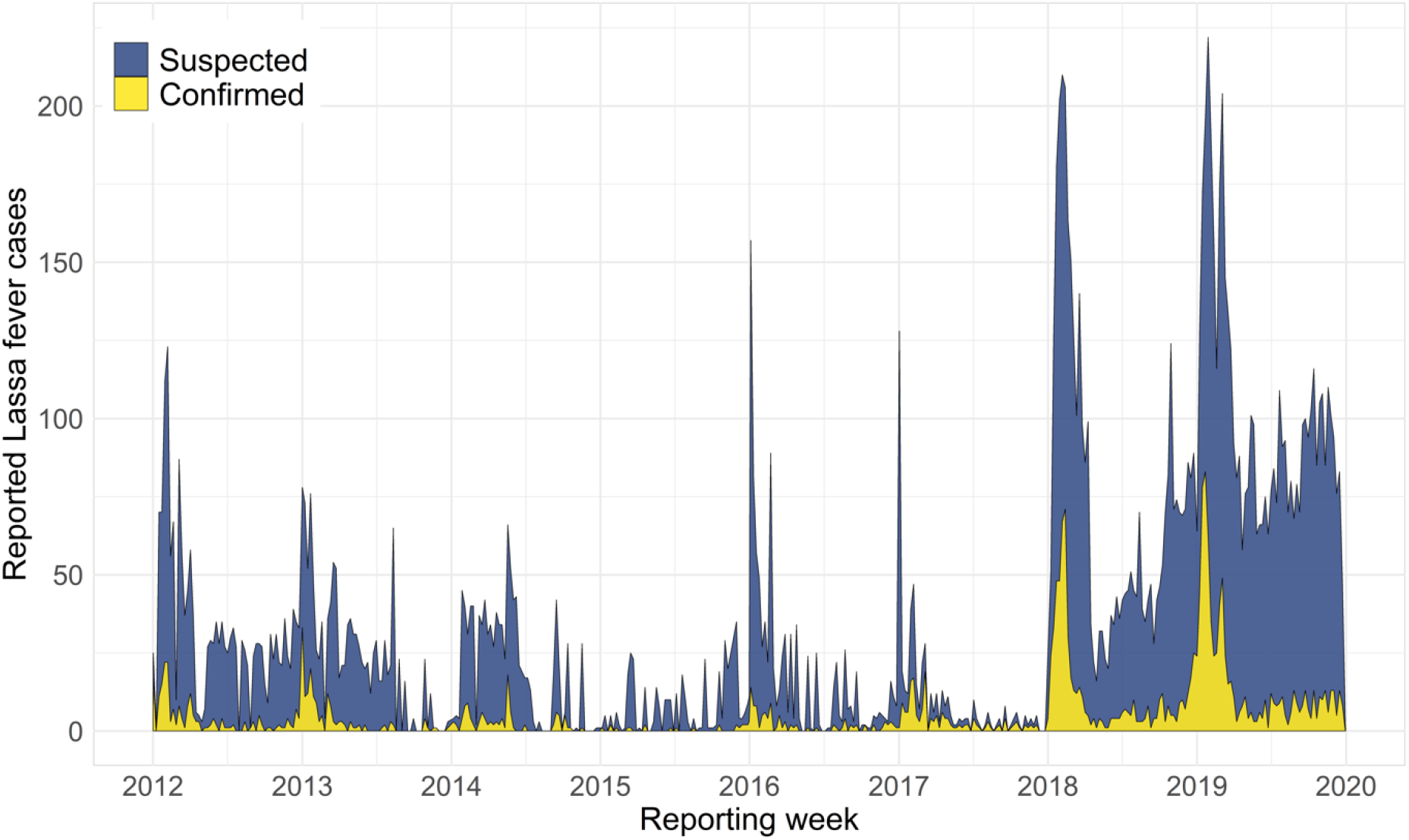
Temporal trends in country-wide Lassa fever case reporting from 2012 to 2019. Polygon height shows the weekly total cases reported across Nigeria, with colour denoting the proportion of cases that were laboratory-confirmed (yellow) or suspected (blue). The full case time series was compiled from two reporting regimes at the Nigeria Centre for Disease Control: Weekly Epidemiological Reports 2012 to 2016, and Lassa Fever Technical Working Group Situation Reports 2017 to 2019 (with case reports followed-up to ensure more accurate counts; datasets shown separately in Extended Data Figure 1). Full description of case definitions and reporting protocols is provided in Methods.

Improvements to country-wide surveillance could, however, be driving any apparent increase in both the incidence and geographical extent of LF in Nigeria. For instance, during the 2012-2019 monitoring period, within-country LF surveillance and response was strengthened under NCDC co-ordination, with a dedicated NCDC Technical Working Group (LFTWG) established in 2016, the opening of three additional LF diagnostic laboratories in 2017-19 (to a total of five; Figure 2), the ongoing roll-out of country-wide intensive training on LF surveillance, clinical case management and diagnosis (Extended Data Table 1)^21^, and the deployment of a mobile phone-based reporting system to 18 states during 2017^22^ (Methods). The result of these improvements is apparent in the smoother case accumulation curves in 2018-19 than observed previously (Extended Data Figure 1), as well as the notable, marked increase in the geographical extent of LF case reports over time. From 2012 to 2015 most reported cases originated from Esan Central in Edo state, the location of Nigeria’s longest-established LF diagnostic laboratory and treatment centre at Irrua Specialist Teaching Hospital (ISTH)^18,19^ (Figure 2). The geographical extent of suspected and confirmed case reports rapidly expanded across Nigeria from 2016 (Figure 2), with a contemporaneous decline in cases from Esan Central. This process can be seen clearly in LGAs surrounding Esan Central (Figure 2, inset) and may reflect increasingly precise attribution of the true geographical origin of cases.

**Figure 2:**
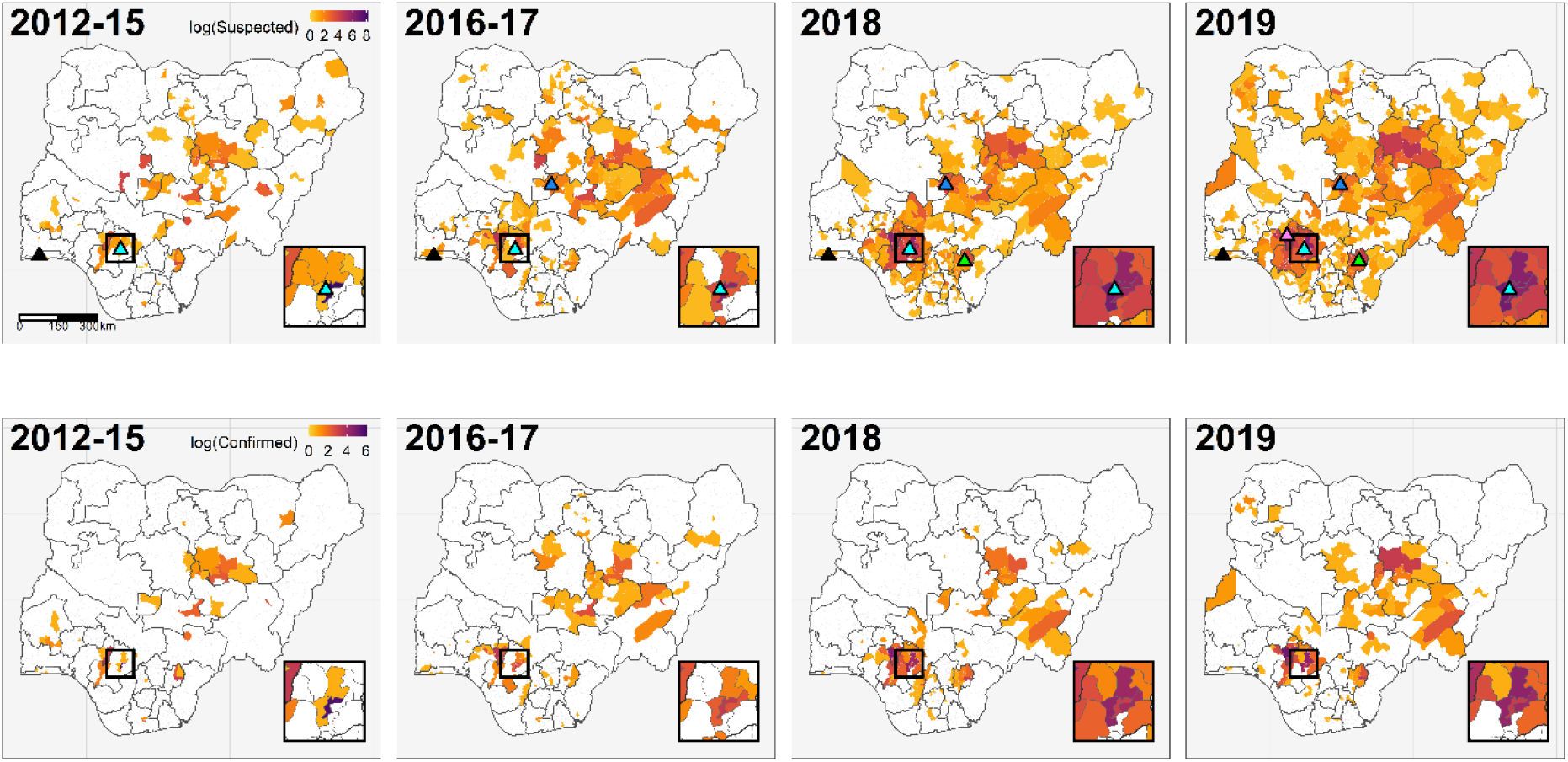
Spatiotemporal trends in Lassa fever surveillance, confirmed cases, and diagnostic laboratory capacity across Nigeria. Maps show, on the natural log scale, the total reported Lassa fever cases (suspected and confirmed; top row) and laboratory-confirmed cases only (bottom row) in each local government authority during the specified year(s). Triangles in the top row show the locations of laboratories with Lassa fever diagnostic capacity. Irrua Specialist Teaching Hospital (Edo, established 2008; inset box, pale blue) and Lagos University Teaching Hospital (Lagos, southwest; black) were both operational since before 2012. Three further laboratories became operational during the study period: Abuja National Reference Laboratory in 2017 (FCT Abuja, north-central; dark blue), Federal Teaching Hospital Abakaliki in 2018 (Ebonyi, southeast; green), and Federal Medical Centre Owo in 2019 (Ondo, south, purple).

To determine the influence of socioecological factors on the geographical distribution of LF risk, we analysed the spatial correlates of annual LF confirmed case occurrence and incidence from 2016 to 2019 inclusive (i.e. the period following the expansion of systematic surveillance) using Bayesian hierarchical models. We adopt a hurdle model-based approach to limit confounding effects of zero-inflation, and separately model annual occurrence across Nigeria (n=774 LGAs across 4 years; binomial likelihood) and incidence-given-presence in LGAs with 1 or more confirmed cases (n=161 across 4 years; case counts modelled as a negative binomial process with log human population as an offset) (Figure 3). In both models we account for country-wide expansion of surveillance using LGA- and State-specific spatially- and temporally-structured random effects, a fixed effect of travel time from nearest laboratory with LF diagnostic capacity, and an adjustment for total annual case reporting effort (incidence model only; Methods, Extended Data Figure 2). Full models with socio-ecological covariates improved model fit compared to random effects-only baseline models and were robust to geographically-structured cross validation tests (Methods, Extended Data Figure 2-3, Supp. Table 1).

**Figure 3:**
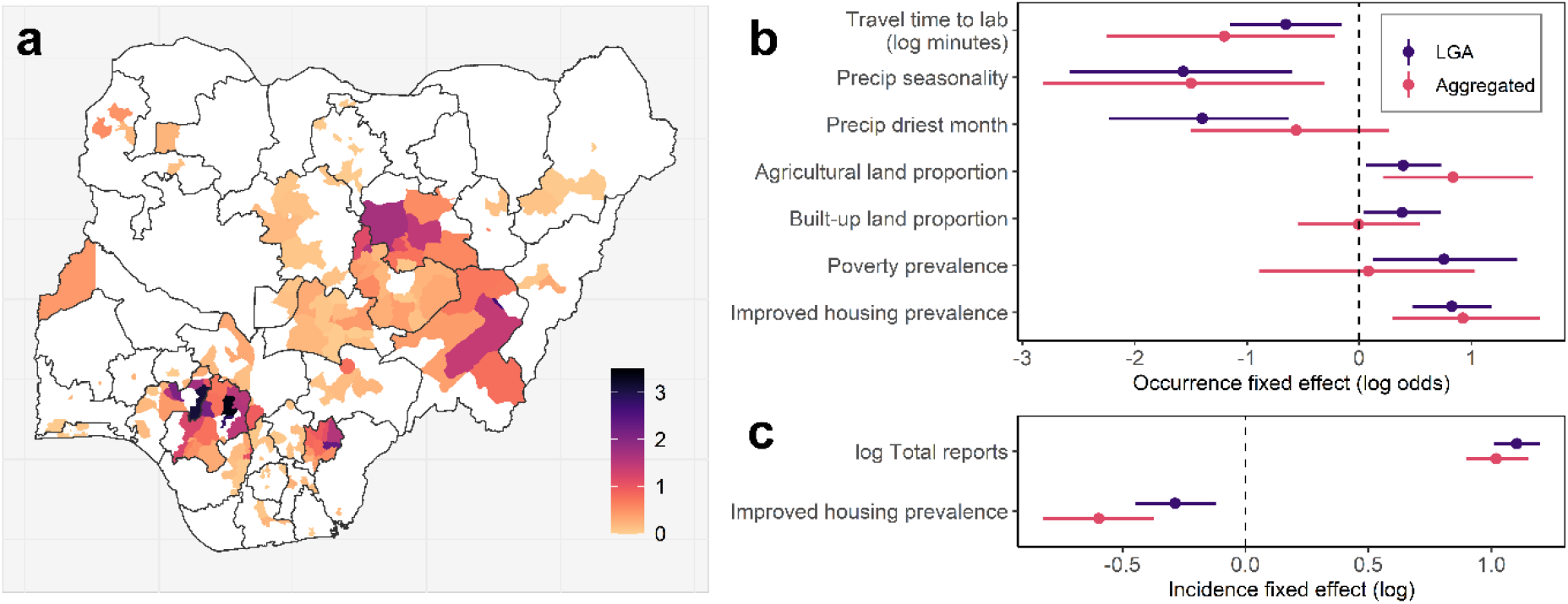
Spatial distribution and correlates of annual Lassa fever occurrence and incidence (2016 to 2019) at local government authority level across Nigeria. Map shows modelled incidence-given-presence (A; cases per 100,000 persons in LGAs with >0 confirmed cases, visualised on the natural log scale), with LGAs with no confirmed cases shown in white. Points and error-bars show socio-ecological fixed effects parameter estimates (posterior mean and 95% credible interval) for best-fitting models of Lassa fever occurrence (B; log odds scale) and incidence-given-presence (C, log scale), fitted to data at either LGA-level (shown in purple) or at lower spatial resolution (130 aggregated districts, shown in pink; Methods, Extended Data Figure 4). Models included spatially and temporally-structured random effects (year, state, LGA), fixed effects for socio-ecological predictors and an adjustment for total reported cases to account for reporting effort (suspected plus confirmed; incidence models only), and were robust to cross-validation tests (Extended Data Figure 3).

We show that LF occurrence is associated with climatic factors (medium-to-high precipitation with fewer seasonal extremes) combined with increasing agricultural land use, which may synergistically affect both reservoir host population sizes and contact with people, as well as several socioeconomic factors that likely jointly influence effective human rodent-contact, healthcare access and LF awareness (Figure 3b; Supp. Table 2). These include strong positive relationships with localised poverty and proximity to LF diagnostic centre, and counter-intuitive positive relationships with urbanisation and improved housing. The latter relationships may be indexing other ecological (e.g. rodent synanthropy), reporting processes (e.g. greater awareness and medical access in urbanised areas) or spatial biases in input data (e.g. new improved housing mainly part of urban encroachment into more natural habitats) not accounted for by other predictors. Overall, the limits of the endemic area of LF appear to be defined, therefore, by the interface of suitable environmental conditions for the reservoir host and socioeconomic conditions that facilitate human-rodent contact, principally rainfed agricultural systems. Notably, after accounting for state-level differences and adjusting for total annual case reporting effort, increasing incidence within non-zero LGAs is predicted solely by reduced prevalence of improved housing (Figure 3c), suggesting that socioeconomic factors regulating human exposure are an important driver of relative risk within environmentally-suitable areas. Public programmes aimed at improving housing and sanitation, and reducing human-rodent contact, may therefore have a positive impact on reducing LF incidence.

Importantly, our results are consistent when modelling at lower spatial resolution (from 774 LGAs into 130 aggregated districts; Extended Data Figure 4), although with weaker or absent effects of urbanisation and poverty on disease occurrence (suggesting these may act on detection or risk at more localised scales; Figure 3b). Spatially projecting the fixed effects of the occurrence model (Extended Data Figure 2) suggests that large contiguous areas of Nigeria are environmentally suitable for LF transmission, and that underreporting may be highest in northern and eastern states. Some localities with high predicted suitability and nearby to existing endemic foci represent key areas to target increased surveillance (e.g. in Oyo, Osun, Ogun and Kogi states). However, LF’s patchy spatial distribution (Figure 3a) and the extremely high incidence in some endemic hotspots (particularly in the south) are mainly explained by reporting-based and random effects, and are poorly predicted by socio-ecological effects alone (Extended Data Figure 2). One interpretation of these results is that the current observed geographical distribution of LF is predominantly shaped by surveillance effort, and that undetected cases are much more ubiquitous than currently recognised; this is plausible given the widespread nature of *M. natalensis*, and indeed is supported by the year-on-year expansion of the endemic area in Nigeria as surveillance continues to be rolled-out^1^. Alternatively, since LASV prevalence in rodents can vary widely over small geographical scales (e.g. between neighbouring villages)^23^, it is also possible that LF risk is highly discontinuous and localised, for unknown reasons. To identify underreported areas and target interventions, therefore, future surveys outside known endemic foci are urgently needed to understand unmeasured social or environmental factors influencing risk (e.g. high public and clinical awareness, agricultural practices^24^ or LASV hyperendemicity in rodents^25,26^).

Understanding and predicting how LF risk dynamics vary seasonally within the currently-identified endemic area is critical to inform disease prevention, control and forecasting. We therefore analysed the spatiotemporal climatic predictors of weekly incidence (2012-2019) in two endemic foci with distinct seasonal climates, agro-ecologies and lineages of LASV^4,27^, in the south (Edo and Ondo; 68% of all reported cases) and north (Bauchi, Plateau and Taraba; 12% of all cases). Within each area we modelled weekly incidence in 5 approximately equal-area districts (i.e. aggregated clusters of LGAs; Methods) as a Poisson process, using district-specific temporally-structured (year and week autoregressive) and unstructured (district) random effects to account for expansions of surveillance, intra-annual seasonality, and baseline differences between districts (Methods). To investigate additional effects of environmental conditions on interannual LF outbreak dynamics and evaluate scope for forecasting, during model selection we considered fixed effects for local precipitation, air temperature and vegetation greenness (Enhanced Vegetation Index; EVI) at 1, 2, 3, 4 and 5 month lags prior to reporting week (to account for delayed effects of climate and delay between infection and reporting; Methods, Extended Data Figure 5).

The full models with lagged climate covariates improved model fit when compared to random effects-only baseline models using several metrics (Methods). We further evaluated the predictive ability of the temporal models using out-of-sample (OOS) cross-validation across the study period, iteratively holding-out 6-month windows, re-fitting the model, and simulating the OOS posterior predictive distribution for the holdout window (Figure 4, Extended Data Figure 6). Models showed good ability to reproduce historical observed case trends in OOS tests (98% of observations falling within 95% predictive interval; Figure 4, Extended Data Figure 7), and climate-driven models reduced root mean square error (RMSE) in OOS predictions relative to baseline models (Supp. Table 1). Separately examining the marginal effects of year, season and climate covariates over time shows that combinations of interannual changes in reporting, natural seasonality, and climatic factors, are sufficient to explain both LF periodicity (distance between peaks) and trends (relative height of peaks over time) in both regions (Extended Data Figure 6). Climatic variability is associated with interannual differences in predicted timing and amplitude of LF seasonality, although notably, conditions during the large case surges in 2018-2019 were within a similar range as in earlier years. This suggests that these unprecedented surges likely resulted mainly from a step change in surveillance and/or other unmeasured factors (Extended Data Figure 6).

**Figure 4:**
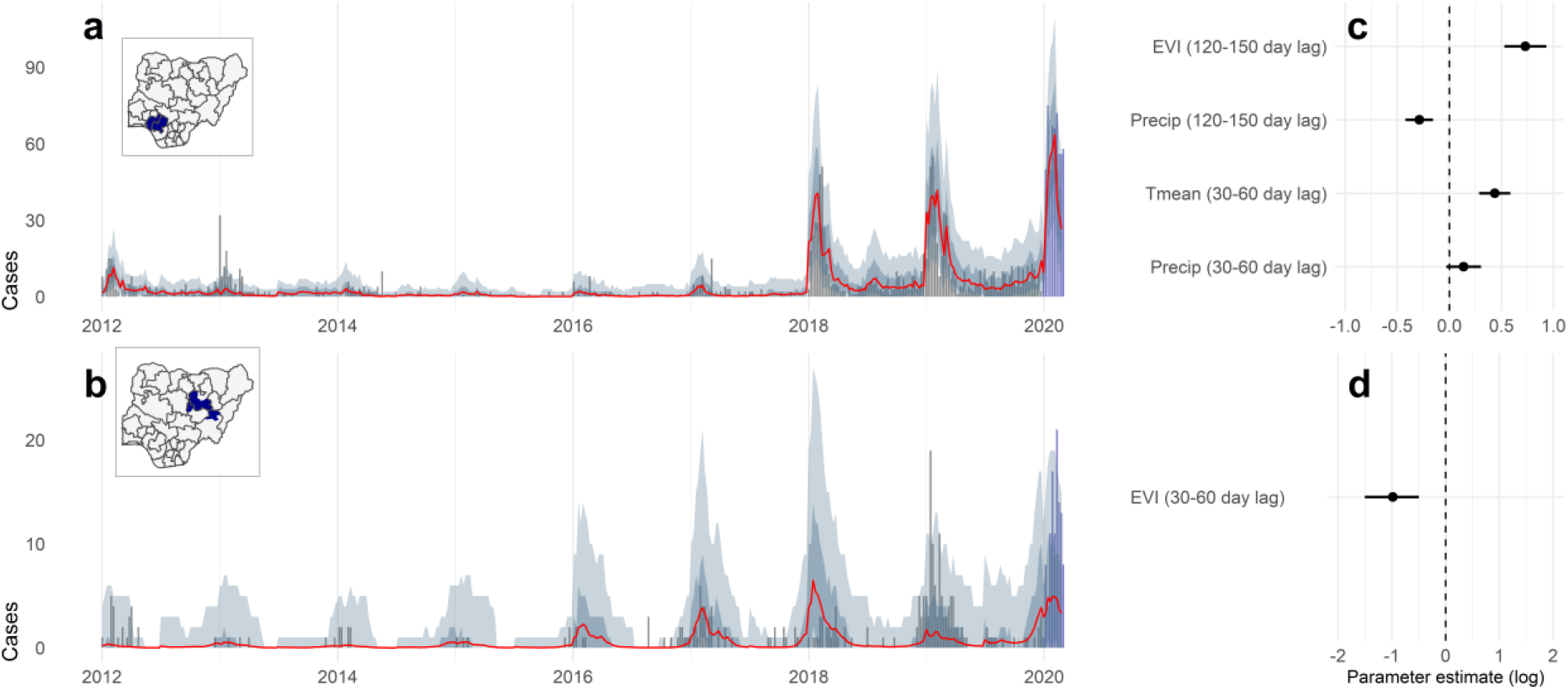
Modelled temporal dynamics and drivers of confirmed Lassa fever cases in south and north Nigeria. Case time series show observed and out-of-sample (OOS) predicted weekly case counts, summed across all aggregate districts in the southern (A; Edo and Ondo states) and northern (B; Bauchi, Plateau and Taraba states) endemic areas. Time series graphs (A-B) show observed counts from 2012 to 2019 (grey bars), OOS posterior median (red line) and OOS 67% (dark grey shading) and 95% (light grey shading) posterior predictive intervals. OOS predictions were made within sequential 6-month windows across the full time series at district-level (i.e. observation-level; Extended Data Figure 7). Dark blue bars show preliminary 2020 weekly case counts for comparison (not included in model fitting), and predictions for 2020 assume reporting effects at 2019 levels (Methods). Inset maps show districts included in the models. The marginal contributions of yearly, seasonal and climatic effects are visualised separately in Extended Data Figure 6. Model fixed effects for precipitation, EVI and daily mean air temperature (Tmean) are shown on the linear predictor (log) scale, separately for southern (C) and northern (D) districts.

In the south, relative increases in LF incidence from the expected seasonal baseline are associated with higher vegetation and lower precipitation at 120-150 days prior to reporting, and higher temperature and precipitation at 30-60 days prior (Figure 4, Supp. Table 3). In the north, we find a strong positive effect of declines in vegetation 30-60 days prior to reporting, although notably including climate information provides smaller improvements in predictive ability than in the south (possibly due to the much lower interannual climate variability during the study period; Extended Data Figure 5). Taken together, these results point towards an important role of climate in explaining LF occurrence and incidence patterns. The effects of lagged rainfall and vegetation dynamics across two agro-ecologically different regions of Nigeria strongly suggests an important role of reservoir host population ecology. Indeed, rainy season characteristics are predictive of subsequent *M. natalensis* population surges and crop damage in East Africa^28^. Temporal variation in rodent and human LASV infections may be driven by seasonal and interannual rodent population dynamics (putatively linked to climate-driven cycles in resource availability and land use^29^) or human agricultural and food storage practices^12^, all of which are important targets for future research. For example, declines in vegetation during the weeks preceding transmission could lead to synchronous food-seeking behaviour in rodents (as natural food availability reduces) and human behaviour changes relating to harvest and crop processing.

The predictive accuracy of these seasonal models (Figure 4; Extended Data Figure 7) suggests that, provided reporting effort is adequately accounted for, lagged climate variables could assist in advance forecasting of LF peaks a month in advance within known endemic areas^30^. To examine this further, we used our models to simulate predicted incidence beyond our study period (to 1^st^ March 2020), fixing all effects except climatic predictors at 2019 levels (i.e. assuming that reporting effort and other interannual differences stay the same). Visual comparison with preliminary state-level case counts from 2020 shows that our climate-driven models successfully predict the higher peaks in 2020, with most weekly counts falling within the predictive interval, particularly in the southern endemic area (Figure 4; Extended Data Figure 8). Notably, our models tend to underpredict 2020 observed case numbers in northern states (Plateau and Taraba), which may be a consequence of recent further improvements in surveillance sensitivity in these areas, or other unobserved events (e.g. hospital-acquired infections). Together with the good OOS predictive performance across the historical time-series, particularly in the highest-incidence districts (Extended Data Figure 6), these findings suggest that a climate-driven approach could in future provide the basis for developing a forecast system in LF-endemic areas. Such early-warning approaches are increasingly used for vector-borne disease control (e.g. dengue^31^) but are rare for directly-transmitted zoonoses. Our dataset is a relatively short time series (n=8 years) and improving these models year-on-year with new surveillance data, and including more precise information on agricultural practices and spatiotemporal variation in land cover, should assist in further clarifying predictors of large case surges and improving forecast accuracy. Our findings have implications for disease control and targeting LASV surveillance in rodents and humans to environmentally-suitable areas where LF is apparently absent, and demonstrate the critical role of improvements in systematic human case surveillance across West Africa^19,20^, as setting the basis for future control of this high priority disease.

## Data Availability

All data will be made available in an online repository upon publication acceptance.

## Methods

### Lassa fever human case surveillance data

We analyse weekly reported counts of suspected and confirmed human cases and deaths attributed to Lassa fever (LF) (as defined in Extended Data Table 1), between 1^st^ January 2012 and 30th December 2019, from across the entire of Nigeria. The weekly counts were reported from 774 local government authorities (LGAs) in 36 Federal states and the Federal Capital Territory, under Integrated Disease Surveillance and Response (IDSR) protocols, and collated by the Nigeria Centre for Disease Control (NCDC). All suspected cases, confirmed cases and deaths from notifiable infectious diseases (including viral haemorrhagic fevers; VHFs) are reported weekly to the LGA Disease Surveillance and Notification Officer (DSNO) and State Epidemiologist (SE). IDSR routine data on priority diseases are collected from inpatient and outpatient registers in health facilities, and forwarded to each LGA’s DSNO using SMS or paper form. Subsequently, individual LGA DSNOs collate and forward the data to their respective State Epidemiologist, also by SMS and paper form, for weekly and monthly reporting respectively to NCDC. From mid-2017 onwards, data entry in 18 states has been conducted using a mobile-phone based electronic reporting system called mSERS, with the data entered using a customised Excel spreadsheet that is used to manually key into NCDC-compatible spreadsheets. Data from this surveillance regime (*‘Weekly Epidemiological Reports’*; WER) were collated by epidemiologists at NCDC throughout the period 2012 to March 2018 (Extended Data Figure 1).

Throughout the study period, within-country LF surveillance and response has been strengthened under NCDC coordination^2,19,32^. LGAs are now required to notify immediately any suspected case to the state level, which in turn reports to NCDC within 24 hours, and also sends a cumulative weekly report of all reported cases. A dedicated, multi-sectoral NCDC LF Technical Working Group (LFTWG) was set up in 2016 with the responsibility of coordinating all LF preparedness and response activities across states. Further capacity building occurred in 2017 to 2019, with the opening of three additional LF diagnostic laboratories in Abuja (Federal Capital Territory), Abakaliki (Ebonyi state) and Owo (Ondo state) (to a total of five; Figure 2) and the roll-out of intensive country-wide training on surveillance, clinical case management and diagnosis. We note that, due to the rapid expansion in test capacity, the definition of a suspected case in our data has subtly changed over the surveillance period: from 2012 to 2016, suspected cases include probable cases that were not lab-tested, whereas from 2017 to 2019, all suspected cases were tested and confirmed to be negative.

In addition to the Weekly Epidemiological Reports data, since 2017 LF case reporting data has also been collated by the LF TWG and used to inform the weekly NCDC LF Situation Reports (*SitRep data*; https://ncdc.gov.ng/diseases/sitreps). This regime includes post-hoc follow-ups to ensure more accurate case counts, so our analyses use WER-derived case data from 2012 to 2016, and SitRep-derived case data from 2017 to 2019 (see Figure 1 for full time series). A visual comparison of the data from each separate time series, including the overlap period (2017 to March 2018) is provided in Extended Data Figure 1, and all statistical models considered random intercepts for the different surveillance regimes. Where other studies of recent Nigeria LF incidence have been more spatially and temporally restricted^33,34^, the extended monitoring period and fine spatial granularity of these data provide the opportunity for a detailed empirical perspective on the local drivers of LF at country-wide scale, and their relationship to changes in reporting effort.

### Data summaries, mapping and processing

We visualised temporal and seasonal trends in suspected and confirmed LF cases within and between years, for both surveillance datasets. Weekly case counts were aggregated to country-level and visualised as both annual case accumulation curves, and aggregated weekly case totals (Figure 1, Extended Data Figure 1). We also mapped annual counts of suspected and confirmed cases across Nigeria at the LGA-level to examine spatial changes in reporting over the surveillance period (Figure 2).

Since analyses of aggregated district data are sensitive to differences in scale and shape of aggregation (the modifiable areal unit problem; MAUP^35^), we also aggregated all LGAs across Nigeria into 130 composite districts with a more even distribution of geographical areas, using distance-based hierarchical clustering on LGA centroids (implemented using ‘hclust’ in R), with the constraint that each new cluster must contain only LGAs from within the same state (to preserve potentially important state-level differences in surveillance regime). Weekly and annual suspected and confirmed LF case totals were then calculated for each aggregated district. We used these spatially aggregated districts to both test for the effects of scale on spatial drivers of LF occurrence and incidence, and as the spatial unit for modelling the seasonal climatic and environmental drivers of LF incidence in endemic districts (see ‘*Statistical analysis’*).

### Statistical analysis

We analyse the full case time series (Figure 1) to characterise the spatiotemporal incidence and drivers of Lassa fever in Nigeria, while controlling for year-on-year increases and expansions of surveillance effort. We firstly modelled examined annual LF occurrence incidence at country-wide scale, to identify the spatial, socio-ecological correlates of disease risk across Nigeria. Secondly, we modelled seasonal and temporal trends in weekly LF incidence within hyperendemic areas in the north and south of Nigeria, to identify the seasonal climatic conditions associated with LF risk dynamics and evaluate scope for forecasting. All data processing was conducted in R v.3.4.1 with the packages R-INLA^36^, raster^37^ and velox^38^. Statistical modelling was conducted using hierarchical regression in a Bayesian inference framework (Integrated Nested Laplace Approximation (INLA)), which provides fast, stable and accurate posterior approximation for complex, spatially and temporally-structured regression models^36,39^, and has been shown to outperform alternative methods for modelling environmental phenomena with evidence of spatially biased reporting^40^.

#### Climatic and socio-ecological covariates

We collated geospatial data on socio-ecological and climatic factors that are hypothesised to influence either *Mastomys natalensis* distribution and population ecology (rainfall, temperature, vegetation patterns), frequency and mode of human-rodent contact (poverty, improved housing prevalence), both of the above (agricultural and urban land cover), or likelihood of LF reporting (travel time to nearest laboratory with LF diagnostic capacity; travel time to nearest hospital). For each LGA we extracted the mean value for each covariate across the LGA polygon using velox^38^. The full suite of covariates tested across all analyses, data sources and associated hypotheses are described in Supplementary Table 2.

We collated climate data spanning the full monitoring period and up until the date of analysis (July 2011 to March 2020). We obtained daily precipitation rasters for Africa^41^ from the Climate Hazards Infrared Precipitation with Stations (CHIRPS) project; this dataset is based on combining sparse weather station data with satellite observations and interpolation techniques, and is specifically designed to support accurate hydrologic forecasts in areas with poor weather station coverage (such as tropical West Africa)^41^. Air temperature daily minimum and maximum rasters were obtained from NOAA, and were also averaged to calculate daily mean temperature. Enhanced Vegetation Index, a measure of vegetation quality, was obtained from processing 16-day composite layers from NASA (National Aeronautics and Space Administration) (excluding all grid cells with unreliable observations due to cloud cover and linearly interpolating between observations to give daily values; Supp. Table 5). We derived several spatial variables to capture conditions across the full monitoring period (Jan 2012 to Dec 2019): mean precipitation of the driest annual month, mean precipitation of the wettest annual month, precipitation seasonality (coefficient of variation), annual mean air temperature, air temperature seasonality, annual mean EVI and EVI seasonality. We also calculated monthly total precipitation, average daily mean (Tmean), minimum (Tmin) and maximum (Tmax) temperature and EVI variables at sequential time-lags prior to reporting week for seasonal modelling (described below in *‘Temporal drivers’*).

We accessed annual human population rasters at 100m resolution from WorldPop (ref). We accessed proportion of the population living in poverty in 2010 (< $1.25 threshold) from WorldPop, to proxy for ability to access risk prophylaxis schemes (e.g. food storage boxes) and for potential susceptibility to disease as a consequence of lower nutrition and co-infection. We accessed proportion of the population living in improved housing in 2015^42^, to proxy for the potential for homes to be infested with rodents. We accessed data on agricultural and urban land cover (proportion of LGA area) for 2015 from processing ESA-CCI rasters.

Finally, we used a global travel friction surface and a least-cost path algorithm^43^ to calculate LGA-level mean travel time to the nearest LF diagnostic laboratory (as a proxy for likelihood of sample testing for LASV) and nearest hospital^44^ (to proxy for probability of patients accessing healthcare when unwell).We acknowledge that such distance-based metrics are coarse approximations of complex processes and subject to limitations. For example, differences in access to transport infrastructure and political unrest will have different effects on reporting in different areas of Nigeria, regardless of proximity to medical facilities, and clinical suspicion for LF will also be influenced by staff training and sensitisation.

Furthermore, diagnostic centres are often established in areas where the disease is already recognised to occur (e.g. in Owo in 2019; Figure 2), so the direction of causality is unclear. The ongoing rollout of electronic reporting systems should in the coming years provide extra information on the role of reporting in determining LF case patterns.

#### Identifying the spatial socio-ecological correlates of annual Lassa fever occurrence and incidence

We modelled annual LF occurrence and incidence at country-wide scale to determine the spatial, socio-ecological correlates of disease across Nigeria. We used annual confirmed case counts per-LGA across the last 4 years of surveillance (2016 to 2019) as a measure of LF incidence, since these years followed the establishment of updated systematic surveillance protocols and the associated geographical expansion of suspected case reports (Figure 2), and so are likely to more fully represent the true underlying distribution of LF across Nigeria. In total, 161 LGAs reported confirmed LF cases from 2016 to 2019, with the majority of cases reported from a much smaller subset (75% from 18 LGAs), and 613 LGAs reported no confirmed LF cases (total=774 LGAs; median 0 cases, mean 2.21, range 0— 321). This overdispersed and zero-inflated distribution presents a challenge for fitting to incidence counts, so we instead adopt a two-stage hurdle model-based approach, and separately model LF occurrence in all LGAs (presence-absence) and incidence-given-presence (i.e. within LGAs that have reported at least 1 confirmed case within the time period, n=161). This both ensures that fitted models adhere to distributional assumptions, and also enables a clearer separation of the contributions of different socio-ecological factors to disease occurrence (i.e. the presence of LF) and to total case numbers in endemic areas.

We model annual occurrence of LF (n=774 LGAs over 4 years) where *y*_*i,t*_ is the binomial presence or absence of LF in LGA *i* during year *t*, and *p*_*i,t*_ denotes the expected probability of LF occurrence, such that:

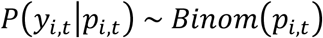

We model annual LF case counts (*z*_*i,t*_) in LGAs with >1 detected case (i.e. non-zero LGAs; n=161 LGAs over 4 years) as a Poisson process, where *μ*_*i,t*_ is the expected number of cases in LGA *i* during year *t*, such that:

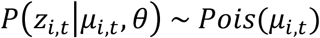

Both *p*_*i,t*_ and *μ*_*i,t*_ are separately modelled as functions of socio-ecological covariates and random effects:

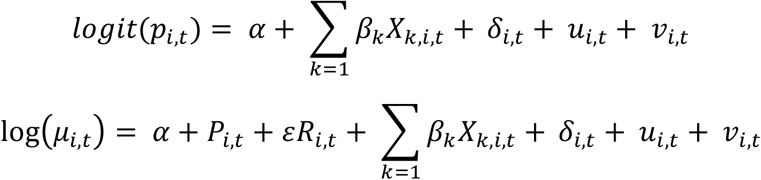

where, for each model, *α* is the intercept; *X* is a matrix of *k* climatic and socio-ecological covariates with *β* linear coefficients; *δ*_*i,t*_ is a structured random effect for year specified at the state-level (first order autoregressive, to account for interannual changes in reporting effort); and LGA-level differences are accounted for using spatially-structured (conditional autoregressive; *v*_*i,t*_) and unstructured i.i.d. (*u*_*i,t*_) random effects jointly specified as a Besag-York-Mollie model. The incidence model additionally includes log human population (*P*_*i,t*_) as an offset, and the logarithm of total case reports (i.e. suspected plus confirmed; *R*_*i,t*_) in LGA *i* and year *t* with coefficient *ε*, to adjust for spatial and seasonal differences in total reporting effort (Figure 2). We set penalised complexity priors for all random effects hyperparameters, and uninformative Gaussian priors for fixed effects.

For both models we considered *β* coefficients for the following covariates: mean precipitation of the driest month, mean precipitation of the wettest month, precipitation seasonality, annual mean air temperature, temperature seasonality, annual mean EVI, EVI seasonality, proportion agricultural land cover, proportion urban land cover, proportion of the population living in poverty (< $1.25 per day), proportion of the population living in improved housing, and two distance-based covariates to account for reporting effort: logarithm of mean travel time to laboratory with LF diagnostic capacity, and logarithm of mean travel time to nearest hospital. We considered both linear and quadratic terms for temperature and rainfall because past studies of *M. natalensis* distribution suggest that these responses may be nonlinear^10,11^. Prior to modelling we removed covariates that were highly collinear with one or more other others (Pearson correlation coefficient >0.8). Continuous covariates not log-transformed were scaled (to mean 0, sd 1) prior to model fitting.

We conducted model inference and selection in R-INLA, and evaluated model fit for both occurrence and incidence models using Deviance Information Criterion (DIC)^45,46^. We conducted model selection on fixed effects by comparing to a random effects-only spatio-temporal baseline model. Covariates were selected for inclusion by removing each in turn from a full model (including all covariates), and excluding any that did not improve fit by a threshold of at least 2 DIC units. The best-fitting models with socio-ecological covariates explained substantially more of the variation in the data relative to baseline models (occurrence ΔDIC=-15.5; incidence ΔDIC=4.4; Supp. Table 1), although most of the spatial heterogeneity in LF in Nigeria was explained by random effects and total case reporting (main text; Extended Data Figure 2). All posterior parameter distributions and residuals were examined for adherence to distributional assumptions (Extended Data Figure 3). We evaluated the sensitivity of spatial model results to geographically-structured cross-validation, in turn fitting separate models holding out all LGAs from each of 12 states that have either high (Bauchi, Ebonyi, Edo, Nasarawa, Ondo, Plateau, Taraba) or low (Kogi, Delta, Kano, Enugu, Imo) documented incidence. Fixed effects direction and magnitude were robust in all hold-out models, indicating that results were not overly driven by data from any one locality (Extended Data Figure 3). We also tested for sensitivity to aggregation scale (i.e. MAUP) by re-fitting the final occurrence and incidence models to the data aggregated into 130 approximately equal-sized districts (as described above). Confirmed LF case totals were calculated for each district, socio-ecological covariates were extracted, and models were fitted as described above (Figure 3b, Extended Data Figure 4, Supp. Table 2).

#### Temporal drivers of Lassa fever cases in high-incidence areas

A growing body of data from clinical records^5,18,20^, ethnographic and social science research^8,48^ and rodent population and serological monitoring^49,50^ suggests that LF risk may be climate-sensitive. Temporal trends in human and rodent infection are hypothesised to be associated with seasonal cycles in rodent population ecology, human land use and food storage practices^12^. We therefore developed spatiotemporal models to quantify the lagged climatic and environmental conditions associated with LF incidence (weekly case counts) across the full duration of surveillance (2012 to 2019). Low and/or variable surveillance effort outside known endemic areas could confound inference of temporal environmental drivers, so here we focus our analyses on regions with case reporting records that span the entire monitoring period. These occur in two main foci: the southern hyperendemic area in Edo and Ondo states, and the northern area spanning Bauchi, Plateau and Taraba states (which in total account for 80% of total confirmed cases since 2012; Figure 3). These areas are distinct in terms of climate, agro-ecology, sociocultural factors and livelihoods^51^, so to avoid potential confounding effects of unmeasured factors, we model the two areas separately using the same protocol (as described below). This approach also provides the benefit of a comparison of temporal environmental correlates in different socio-ecological settings.

Within each area (henceforth referred to as Southern and Northern endemic areas) we fit models to LF time series from five spatially-aggregated districts (clusters of LGAs as derived above): spatial aggregation better harmonises the resolution of disease data with climatic data, and reduces potential noise associated with uncertain attribution of the true LGA of origin for cases in the early part of the time series (especially in Southern states; see Figure 2). In each endemic area, we model weekly case counts *z*_*i,t*_ (n=5 districts over 8 years, so total 2090 observations per model) as a Poisson process:

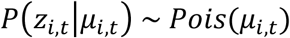

where *μ*_*i,t*_ is the expected number of cases for district *i* during week *t*, modelled as a log-link function of climatic covariates and temporally-structured random effects to account for seasonality and changes in reporting effort:

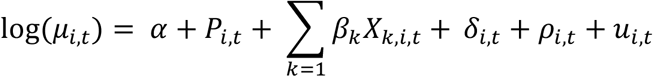

where *α* is the intercept, *P*_*i,t*_ is log human population included as an offset (thereby modelling incidence), *X*_*k,i,t*_ is a matrix of *k* climatic predictor variables with *β* linear coefficients, *δ*_*i,t*_is a district-specific temporally-structured effect of year (first-order autoregressive, to account for ongoing changes in reporting effort and other interannual differences), *ρ*_*i,t*_ is a district-specific seasonal effect of epidemiological week to account for seasonality (first order autoregressive model to capture dependency between weeks), and *u*_*i,t*_ is an unstructured (i.i.d.) random effect to account for additional district-level differences. We set penalised complexity priors for all hyperparameters and uninformative Gaussian priors for fixed effects.

For each endemic area we considered *β* coefficients for 5 lagged climatic variables: mean precipitation, EVI, and mean, minimum and maximum daily air temperature, calculated across a 30-day window at time lags beginning from 30-days to 150-days prior to reporting week (i.e. 30-60, 60-90, 90-120, 120-150 and 150-180 days; spanning 1 to 6 months before reporting). We considered lagged climate variables to account for, firstly, delayed effects of seasonal environmental cycles on *M. natalensis* population ecology, behaviour and LASV prevalence that are hypothesised to influence force of infection to humans^49^, and secondly, delays between LASV infection event, disease incubation period (which can be up to 10 days^12^) and patient presentation at a medical facility. Including climate information at a sufficient lag (1 month upward) also enables us to evaluate the scope for climate-driven predictive forecasting of seasonal LF peaks to support disease control. We did not include the temporally-invariant covariates included in the spatial models, since the smaller number of districts (five per endemic area) provides low comparative power to detect any spatial effects on incidence.

We first a baseline model with seasonal and spatial random effects only. During model selection we selected for the combination of lagged climatic effects that best explain observed weekly incidence trends, again selecting among models using DIC, and here conducting forwards stepwise selection (due to the high number of potentially collinear covariates) with a threshold of 6 DIC units (as we were selecting for significant improvements in predictive ability)^52^. Including climate information explained substantially more of the variation in data relative to baseline models, although with much larger improvements in the south than north (Southern ΔDIC=-74.7, Northern ΔDIC=-7.67 and similar improvements in two other metrics, WAIC and root mean square error, Supp. Table 1). To evaluate predictive ability of the models on unseen data, we then conducted out-of-sample posterior predictive tests using a 6-month holdout window across the full time series. For holdout models we excluded all observations from each sequential 6-month window (Jan-Jun; Jul-Dec), re-fitted the model, drew 2500 parameter samples from the approximated joint posterior distribution, then used these to (1) calculate OOS posterior mean and intervals and (2) simulate the OOS negative binomial predictive distribution (i.e. the range of plausible expected case counts given the model). We calculated and visualised the proportion of observed values falling within 67% and 95% OOS predictive intervals, overall and over time (Extended Data Figure 7). Both climate models improved predictive ability on out-of-sample data relative to baseline models, measured using RMSE (Supp. Table 1) although again with much larger improvements in the south (potentially due to much wider climate variability during the study period; Extended Data Figure 5).

Finally, to evaluate scope for model-based forecasting, we used the climate models to forecast posterior mean and predictive intervals for the 2020 Lassa fever season (using climate data up to March 2020) from the model fitted to the full time series. We compared these predictions to 2020 preliminary state-wide confirmed case counts compiled for the NCDC Situation Reports, holding yearly random effects (*δ*_*i,t*_) at the same level as 2019 (i.e. predictions for 2020 assume the same level of effort; Figure 4, Extended Data Figure 8).

Because these are preliminary data they are unsuitable for model fitting, but provide a useful future OOS test for forecasting ability. In earlier versions of the manuscript and conference presentations developed prior to 2019, we had conducted the same forecasting test for the 2019 Lassa fever season, and successfully predicted a substantially larger 2019 peak than in 2018 when presenting at the NCDC Lassa fever conference in Abuja, Nigeria in January 2019.

## Data availability

All data used for these analyses are provided at the accompanying repository [to be added in]

## Code availability

All code used for these analyses are provided at the accompanying repository [to be added in]

## Acknowledgements

The authors thank all the epidemiologists and clinical staff at Nigeria state and Local Government Authority levels who collected and submitted the original case reports.

## Author contributions

CCD-N, EI, YRU, OHS, AOM, IA, OBI and CI collected the data; DWR, RG, LE, IA, KEJ and CI designed the study, RG led and conducted the analyses with DWR and LE, and RG, DWR and KEJ wrote the manuscript. All authors contributed to writing and editing the full manuscript.

## Materials & Correspondence

All correspondence should be directed to David Redding via the email address

## Funding

This research was supported by an MRC UKRI/Rutherford Fellowship (MR/R02491X/1) and Wellcome Trust Institutional Strategic Support Fund (204841/Z/16/Z) (both DWR), a Graduate Research Scholarship (RG) and Global Engagement Fund grant (DWR, RG) both from University College London, and the Natural Environment Research Council (NERC) PhD studentship (LE). CAD acknowledges joint Centre funding from the UK Medical Research Council and Department for International Development (DFID). CAD is funded by the Department of Health and Social Care using UK Aid funding on a grant managed by the UK National Institute for Health Research (NIHR). The views expressed in this publication are those of the author(s) and not necessarily those of the Department of Health and Social Care. IA acknowledges funding from the UK NIHR (NF-SI-0616–10037), EDCTP PANDORA Consortium and the UK MRC. KEJ acknowledges the Dynamic Drivers of Disease in Africa Consortium, NERC project no. NE-J001570-1 which was funded with support from the Ecosystem Services for Poverty Alleviation Programme (ESPA). The ESPA programme was funded by DFID, the Economic and Social Research Council (ESRC) and NERC.

**Supplementary Information** is available in the online version of the paper.

## Extended Data

**Extended Data Figure 1:**
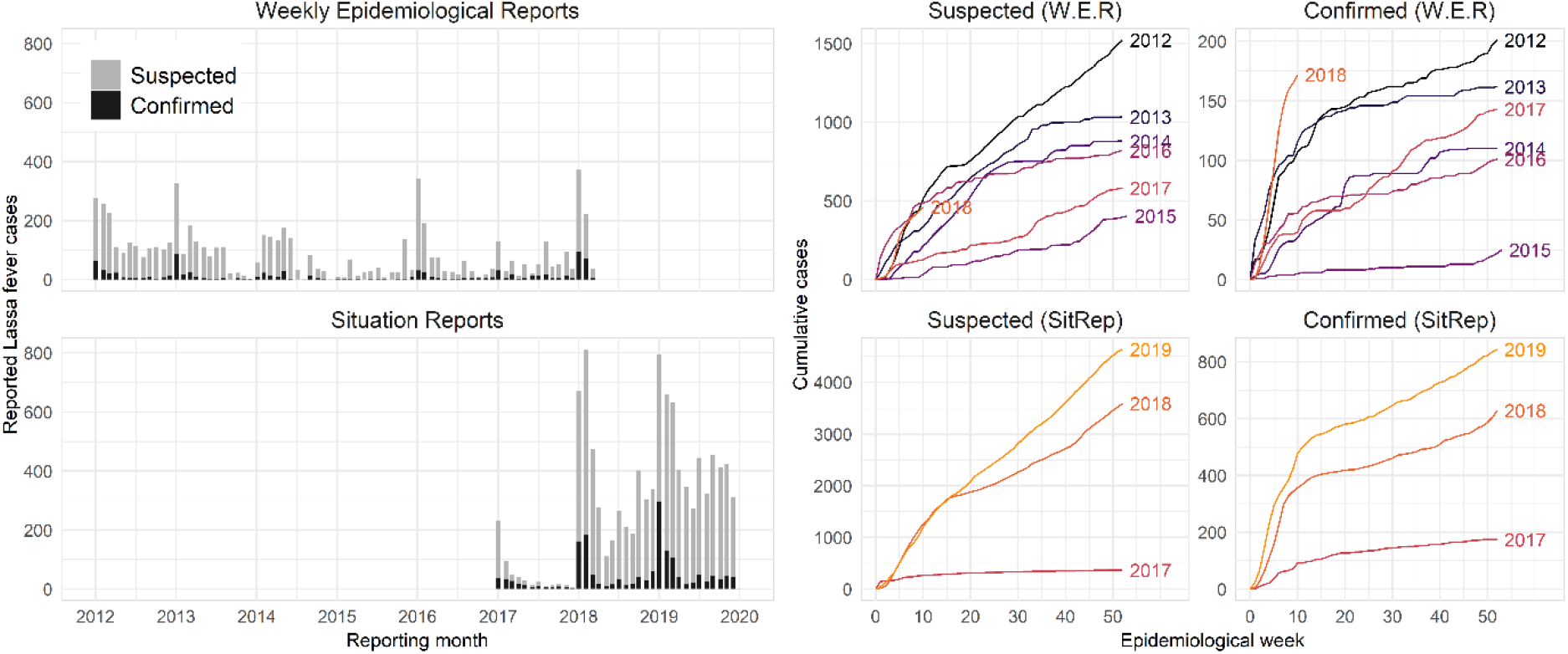
Lassa fever case time series from Nigeria Centre for Disease Control reporting regimes. Graphs summarise weekly surveillance data aggregated across all local government authorities (LGAs), from January 2012 to December 2019, and show the differences between two surveillance regimes. Bar plots show monthly total cases from the long-term Weekly Epidemiological Reports (W.E.R., top) and more recent Situation Reports (SitRep, bottom) regime, with bar heights representing the total LF cases from all epidemiological weeks starting during a given month, split into suspected (grey) and confirmed (black) cases. Weekly case accumulation curves per-surveillance regime, per-year show total reported cases (including both suspected and confirmed; top graphs) and confirmed only (bottom graphs). LF trends during the overlap period between the two regimes are similar (January 2017 to March 2018) but the SitRep data (based on the most current reporting regime and including post-hoc follow-ups to ensure accurate counts) more clearly show the very large increase in both suspected and confirmed case reports in 2018. The full time series used in analyses (Figure 1) includes W.E.R. data from 2012 to 2016, and SitRep data from 2017 to 2019.

**Extended Data Figure 2:**
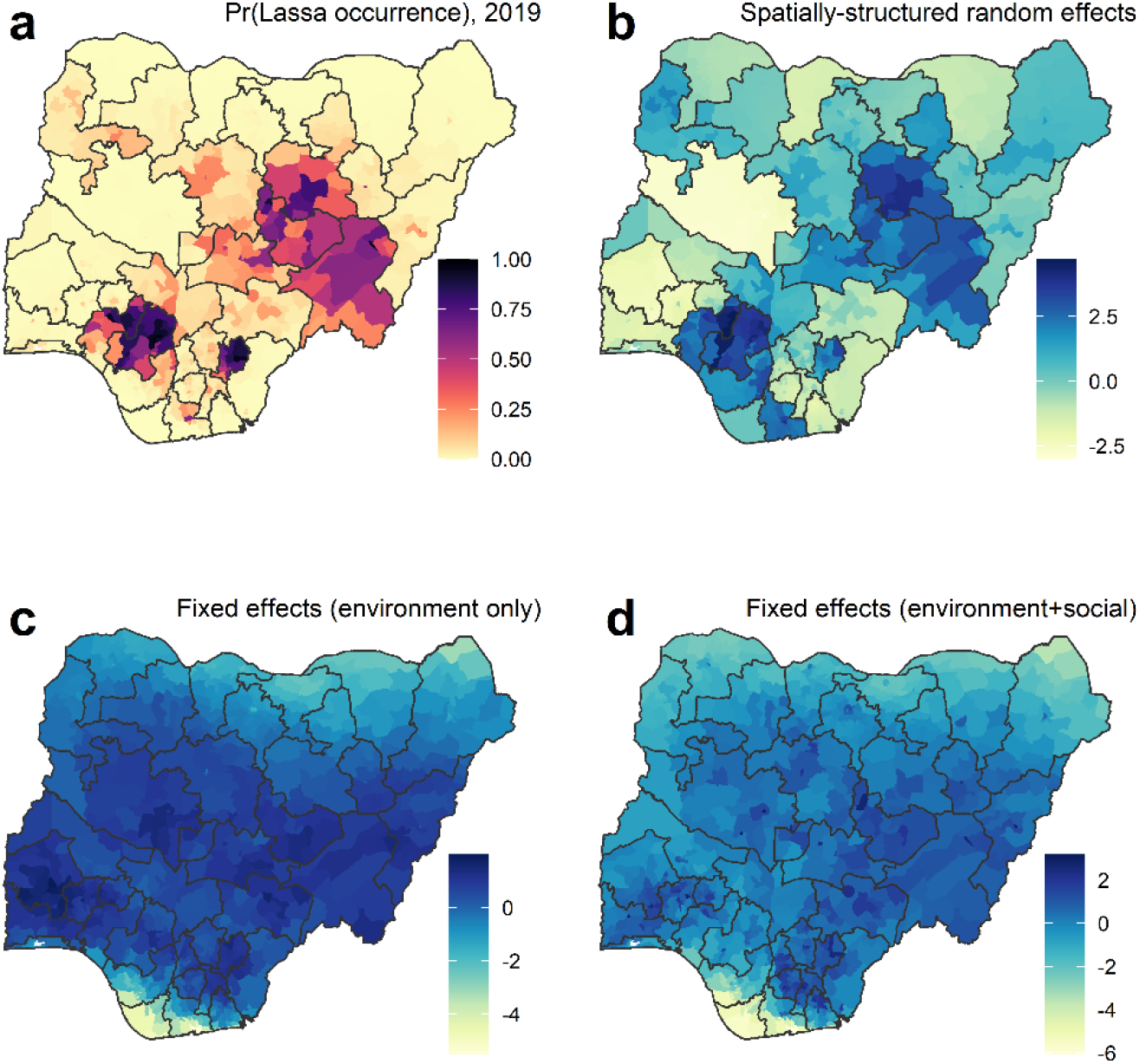
Components of the spatial models of Lassa fever occurrence and incidence across Nigeria. We spatially modelled annual LF occurrence (binomial) and incidence-given-presence (negative binomial, in local government authorities with >0 confirmed cases) from 2016 to 2019 (774 LGAs, 4 years). Model-fitted probability of LF occurrence is shown for 2019 (A). The other plots show marginal effects of occurrence model components on the linear predictor scale (log-odds). Much of the variation in observed occurrence, in particular the southern hyperendemic area (Edo and Ondo states) is explained by spatially and temporally-structured random effects (LGA, year, state; B). Contributions of socio-ecological fixed effects to the linear predictor are projected on the log-odds scale (C-D). Projected environmental (climate and agriculture; E) and combined socioeconomic and environmental effects (C) show that the broad envelope of LF suitability covers much of Nigeria. However, these covariates alone are insufficient to explain either the heterogeneous distribution of reported LF or the high-incidence southern hotspot, which are mainly explained by random effects (B).

**Extended Data Figure 3:**
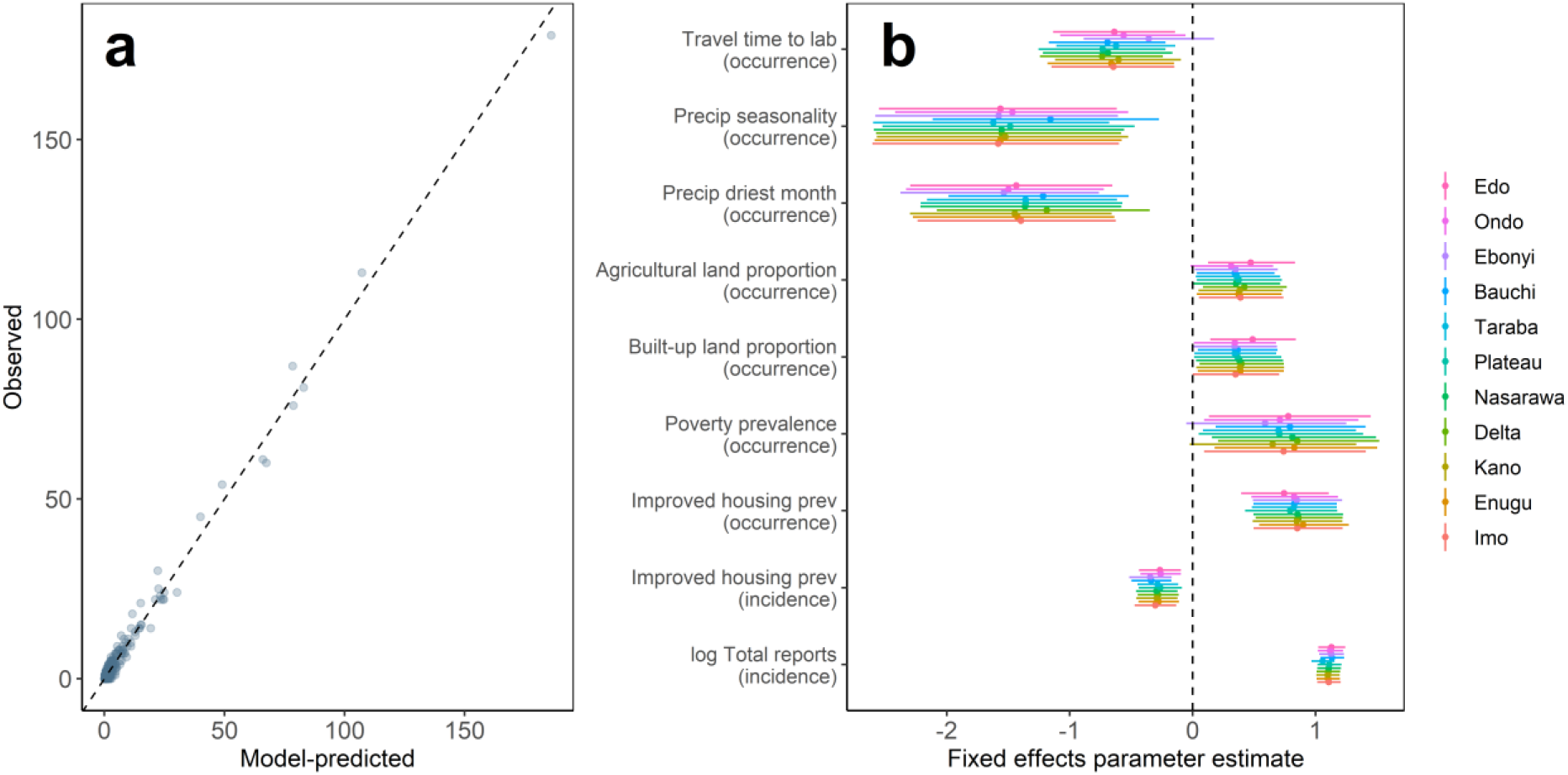
Model fit and geographical cross-validation for spatial models of Lassa fever occurrence and incidence. We selected spatial occurrence and incidence models using model selection on information criteria to determine the best-fitting candidate model (Methods), and the best models with socio-ecological fixed effects showed improved performance over a random effects-only baseline model (occurrence: delta-DIC =; incidence: delta-DIC =). The full incidence model showed close fit to observed case counts (A). The direction and magnitude of fixed-effects (mean ± 95% credible interval) was also robust to geographically-structured cross-validation (B), which involved in turn excluding all LGAs from each of 12 Lassa-endemic and non-endemic states (ordered by total case counts, top to bottom), indicating that the results were not overly influenced by data from any one locality.

**Extended Data Figure 4:**
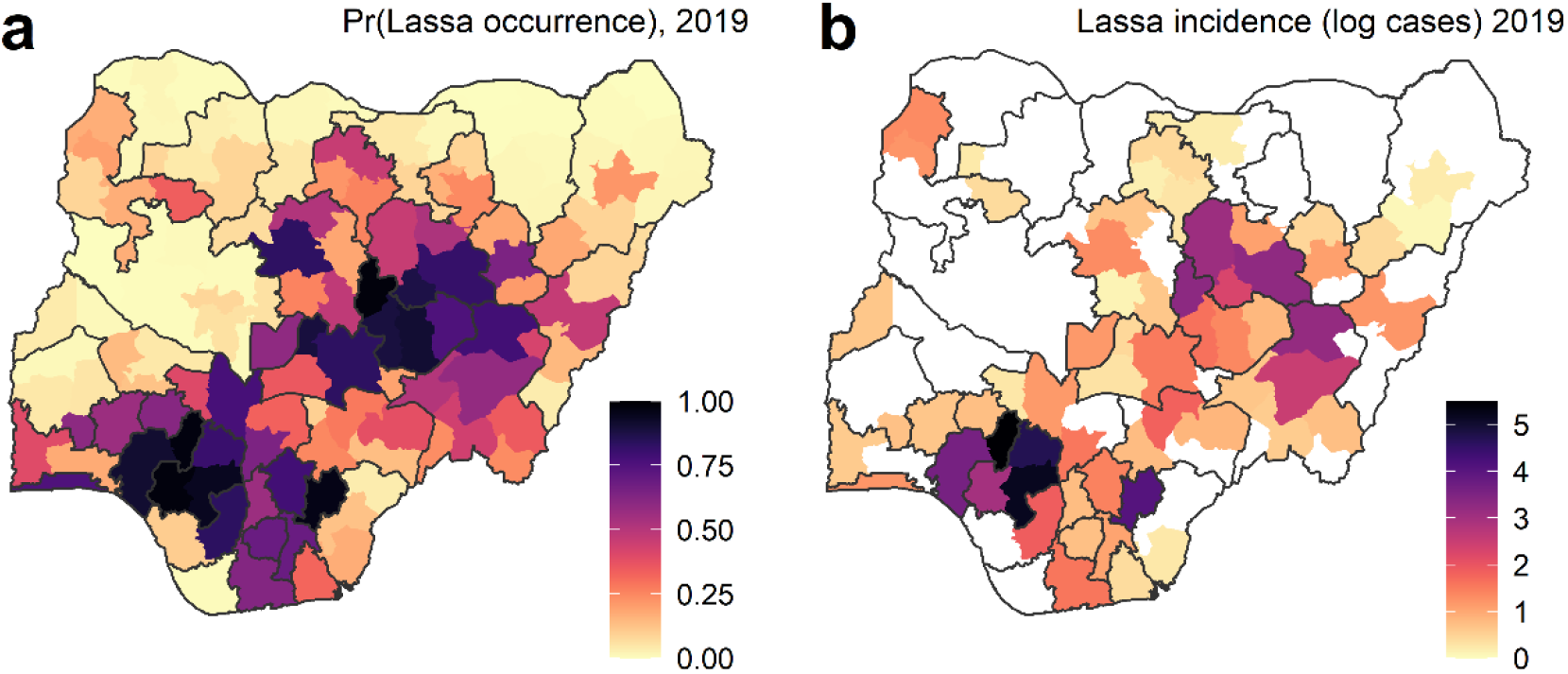
Fitted spatial models of Lassa fever occurrence and incidence for spatially-aggregated districts. Analyses of areal data can be confounded by differences in scale and shape of aggregated districts, and local government authority (LGA) geographical areas in Nigeria are highly skewed and vary over >3 orders of magnitude (median 713km^2^, mean 1175km^2^, range 4 – 11255km^2^). To examine the effects of scale on inferences, we repeated all spatial modelling after aggregating LGAs into 130 composite districts, subject to the constraints of state boundaries, producing a more even area distribution (median 6826km^2^, mean 6998km^2^, range 1641 – 14677km^2^). Maps show fitted values for 2019 from the spatial models of occurrence (A) and incidence-given-presence (B; with districts with no confirmed LF cases shown in white), including the same random and fixed effects structure as the LGA-level model (main text, Figure 3). Model parameter estimates are provided in Supp. Table 2.

**Extended Data Figure 5:**
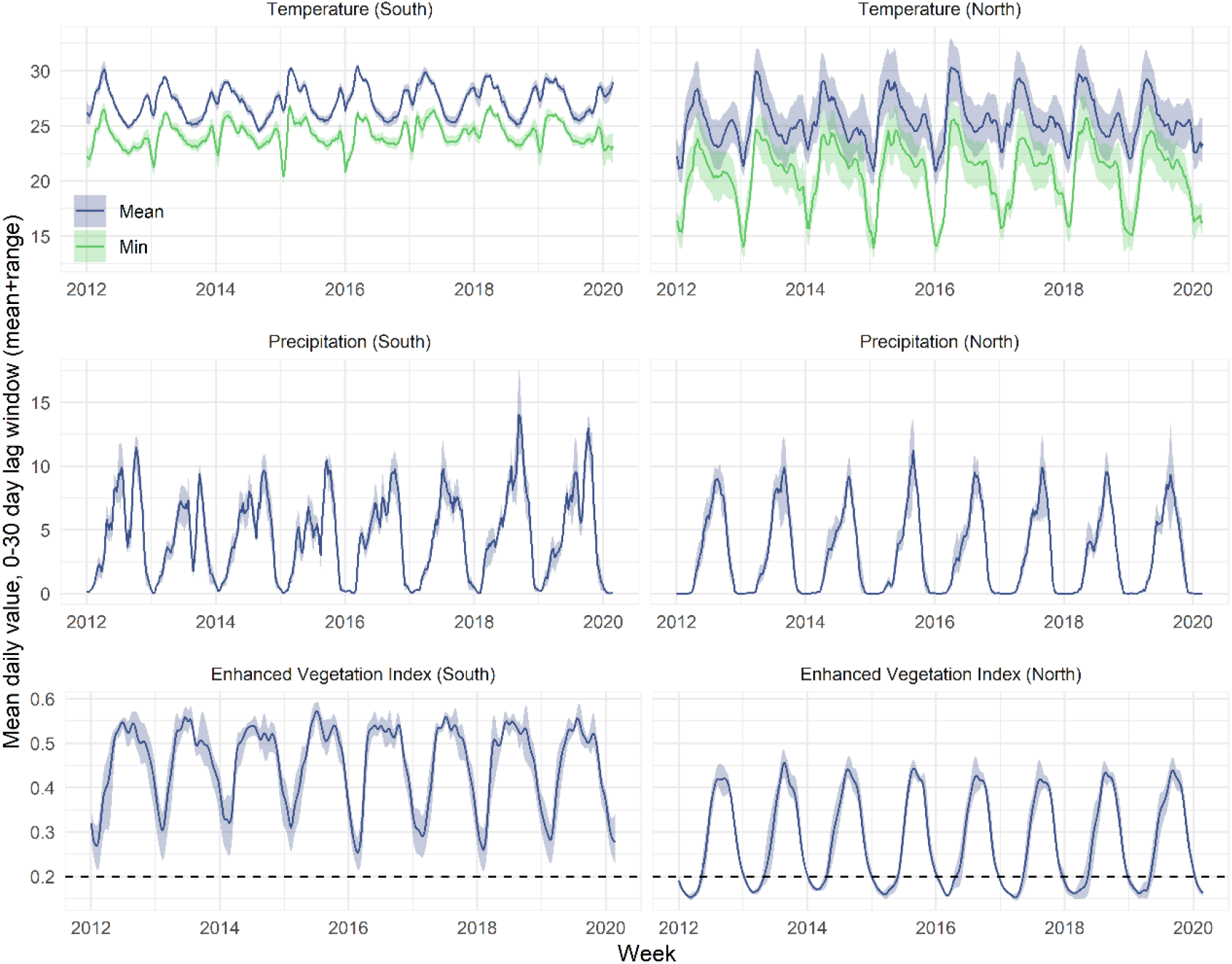
Seasonal and interannual climate and vegetation dynamics in Lassa-endemic regions of south and north Nigeria. Graphs show, for south (left column) and north (right column) Lassa-endemic areas, district-level weekly mean environmental (temperature, precipitation) and vegetation values across a 30-day window prior to reporting week (i.e. at time of transmission occurring). Lines and error-bars show the mean and range of weekly values, across all aggregated districts included in temporal models (5 per region). Temperature estimates are daily mean (Tmean; blue) and minimum (Tmin), derived from Climate Prediction Centre interpolated air temperature layers from NOAA (Tmax excluded because of similarity to Tmean). Precipitation estimates are daily mean rainfall, derived from modelled daily rainfall layers from CHIRPS Africa. Vegetation estimates are daily mean Enhanced Vegetation Index (EVI), derived from processed and linearly interpolated 16-day interval EVI rasters from NASA. EVI values below a ∼0.2 threshold (dotted line) indicate a lack of dense green vegetation.

**Extended Data Figure 6:**
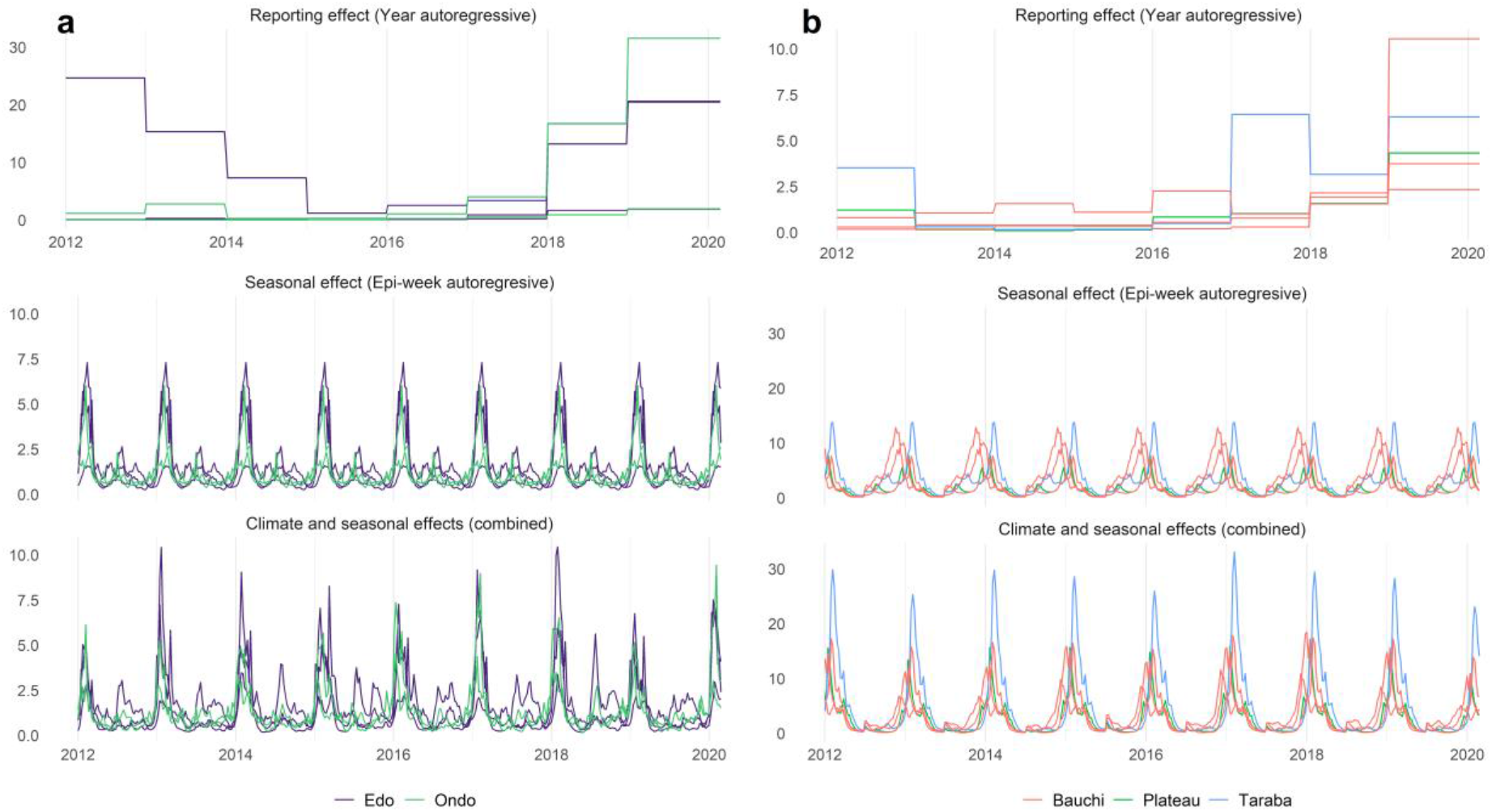
Marginal effects of environment, seasonality and year on temporal Lassa fever incidence. Figures show the mean marginal contributions of different model components to weekly predicted LF incidence in the southern (A) and northern (B) endemic areas of Nigeria. Separate lines are mean combinations of linear effects for each district (5 per model, coloured by state), and are exponentiated to show proportional change in effects on incidence over time, so are not interpretable on the absolute scale. The top row shows the district-specific yearly autoregressive random effect (*δ*_*i,t*_) which reflects between-year differences in reporting effort and potentially other unmeasured factors. From 2012 to 2015, reporting was low in all districts except Esan in the south (the location of Irrua Specialist Teaching Hospital; Figure 2), and reporting across all districts has increased year-on-year since 2016. The middle row shows the district-specific seasonal autoregressive random effect (*ρ*_*i,t*_), which reflects the average intra-annual seasonality of LF across all study years. The bottom row shows the combination of climatic effects and seasonal effects (i.e. ∑_*k*=1_ *β*_*k*_*X*_*k,i,t*_ + *ρ*_*i,t*_), and shows the additional expected contribution of climatic predictors to interannual seasonal differences in LF incidence.

**Extended Data Figure 7:**
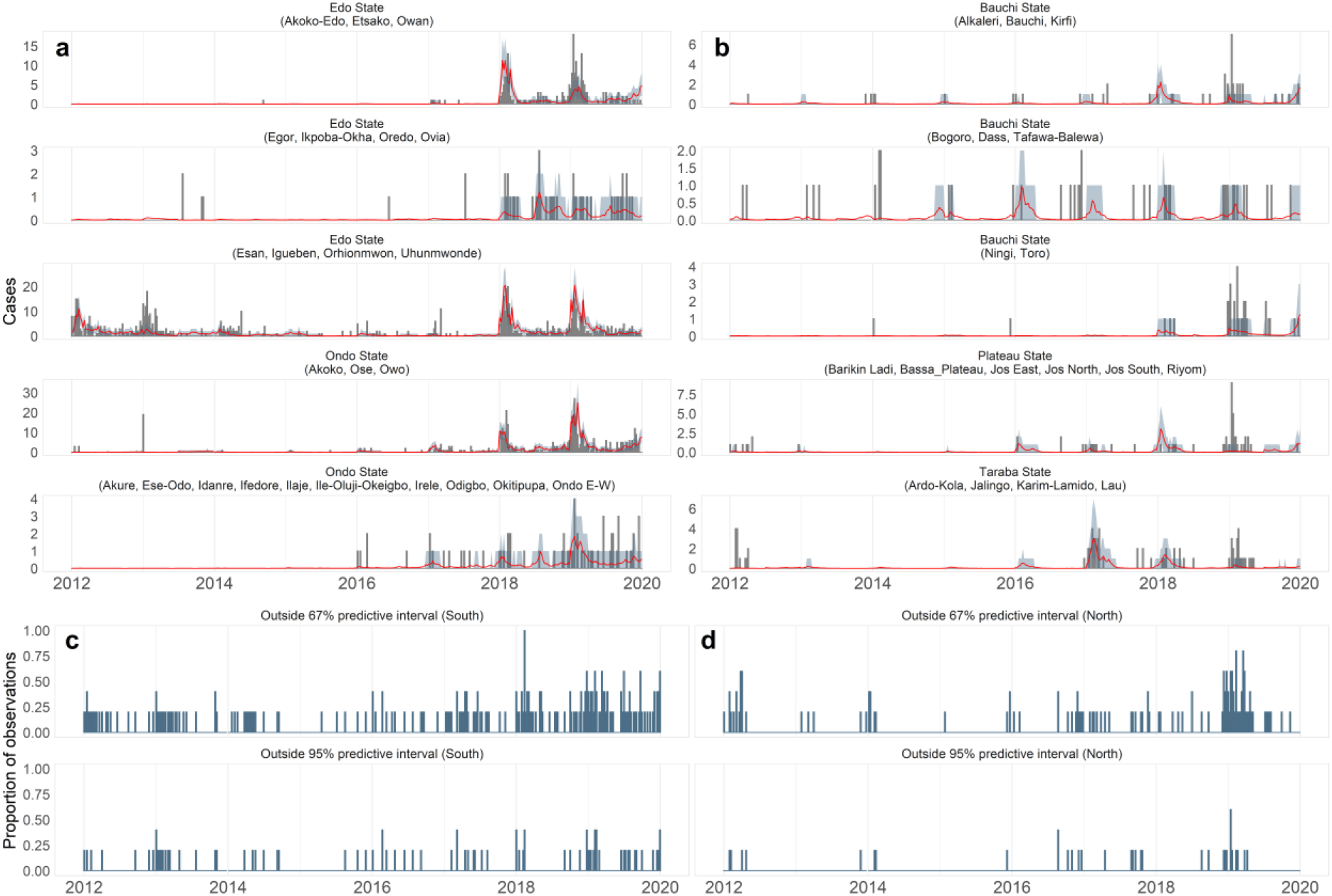
Observation-level out-of-sample trends and posterior predictive distributions for temporal models of Lassa fever incidence. Trend graphs show, for each aggregated district included in models of the Southern (A) and Northern (B) endemic areas, the weekly observed confirmed LF cases (grey bars), out-of-sample (OOS) posterior median and 95% credible interval (red line and ribbon), and OOS 67% and 95% simulated posterior predictive intervals (Methods). Plot names refer to the state, and local government authorities within the district in brackets. Blue barplots show, for South (C) and North (D), the proportion of weekly observations (5 per week) falling outside the 67% (top plot) and 95% (bottom) posterior predictive intervals (i.e. higher proportion indicates lower accuracy). Across the entire surveillance period, predictive accuracy was good for both the Southern (89% of observations falling within the 67% predictive interval; 96% within the 95% interval) and Northern models (96% of observations within 67% interval and 98% within the 95% interval).

**Extended Data Figure 8:**
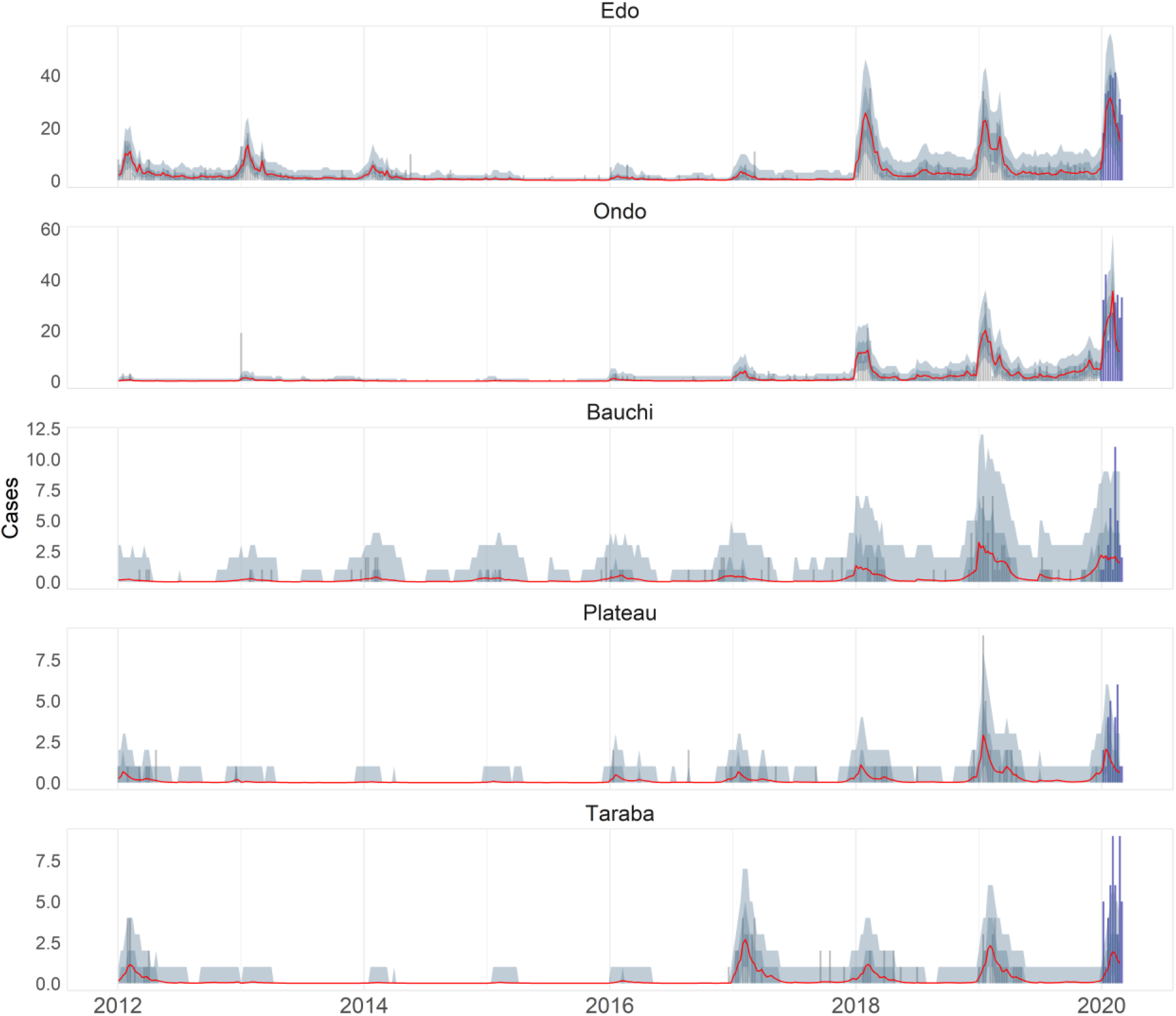
Modelled time series and 2020 forecasts for endemic states. Trend graphs show, for each state included in temporal models, the weekly observed confirmed LF cases (grey bars), posterior median (red line), 67% and 95% simulated posterior predictive intervals (ribbons), and preliminary weekly state-level case counts forecast for 2020 (blue bars; not used to fit the models) (Methods). Predictions were made using separate southern (Edo, Ondo) and northern (Bauchi, Plateau, Taraba) models and 2020 predictions assume 2019 levels of reporting effort. The models in general are successful at predicting the 2020 case surge peak, although one exception is clear underprediction in Taraba state, which may be due to substantially increased state-level reporting effort in 2020^1^, and/or a mismatch between district-level predictions (which are only for the far north of the state; Figure 4) and state-level preliminary counts.

**Extended Data Table 1:** Clinical definitions used for diagnosis of suspected and confirmed Lassa fever cases. Clinical definitions and criteria are listed in weekly Nigeria Centre for Disease Control Lassa fever Situation Reports^1^.

| Term | Primary Criterion | Secondary Criterion |
| --- | --- | --- |
| Alert case | Any person who has an unexplained fever (i.e. Malaria and other likely causes of fever have been ruled out), with or without bleeding<br><u>OR</u><br>Any person who died after an unexplained severe illness with fever and bleeding |  |
| Suspected case | An illness of gradual onset with one or more of the following: malaise, fever, headache, sore throat, cough, nausea, vomiting, diarrhoea, myalgia (muscle pain), central chest pain or retrosternal pain, hearing loss and either: | **a.** History of contact with excreta or urine of rodents<br><u>OR</u><br>**b.** History of contact with a probable or confirmed Lassa fever case within a period of 21 days of onset of symptoms<br><u>OR</u><br>Any person with inexplicable bleeding/haemorrhaging |
| Confirmed case | Any suspected case with laboratory confirmation (PCR is the key diagnostic tool used here). |  |
| Deaths | Cases that die either as suspected or confirmed cases. |  |
| Alert threshold | A single suspected case of Lassa fever. |  |
| Outbreak threshold | A single confirmed case of Lassa fever. Clinicians should have a high index of suspicion when managing febrile illnesses; especially cases with: | **a.** A history of non-response to antimalarials or antibiotics.<br>**b.** A compatible history of travel to an endemic area or an area with an ongoing outbreak, contact with a confirmed case of Lassa fever, negative thick blood film for malaria parasite are suggestive.<br>**c.** Signs of haemorrhage and shock which is strongly suggestive, but these signs often appear late in the illness. |

## Supplementary Information

**Supplementary Table 1:** Metrics of model fit and predictive ability for spatial and temporal Lassa fever models. Tables show the comparison of measures of model fit (Deviance Information Criterion DIC; Watanabe-Akaike Information Criterion WAIC; root mean square error RMSE) between baseline random-effects only models, and models with socio-ecological and climate covariates, for the spatial models (A) and temporal incidence models (B). For the temporal models WAIC, DIC and RMSE are calculated for the models fitted to the full dataset (within-sample), and RMSE is also calculated out-of-sample for the full time-series, and for the later high-incidence years (based on OOS cross-validation, iteratively fitting the model with sequential 6-month windows held out).

A: Spatial models of occurrence and incidence
| Response | Model | DIC | WAIC | Socio-ecological covariates |
| --- | --- | --- | --- | --- |
| Occurrence | Baseline | 1195.4 | 1188.9 | None |
| Occurrence | Socio-ecological | 1179.3 | 1178.9 | Travel time to Lassa lab, Precip seasonality, Mean precip of the driest month, Proportion urban, Proportion agricultural, Poverty prevalence, Improved housing prevalence |
| Incidence | Baseline | 1341.4 | 1338.2 | None |
| Incidence | Socio-ecological | 1337.3 | 1335.1 | Improved housing prevalence |

B: Temporal models of incidence in endemic areas
| Region | Model | DIC | WAIC | RMSE (within-sample) | RMSE (out-of-sample, all years) | RMSE (out-of-sample, 2018-2019) | Climate covariates |
| --- | --- | --- | --- | --- | --- | --- | --- |
| South | Baseline | 3054.6 | 3180.9 | 1.235 | 1.827 | 2.965 | None |
| South | Climatic | 2979.9 | 3093.8 | 1.135 | 1.688 | 2.688 | Tmean (30-60d), EVI (120-150d), Precip (30-60d), Precip (150-180d) |
| North | Baseline | 1177.7 | 1197.4 | 0.382 | 0.511 | 0.85 | None |
| North | Climatic | 1170.6 | 1187.9 | 0.382 | 0.503 | 0.836 | EVI (30-60d) |

**Supplementary Table 2:**
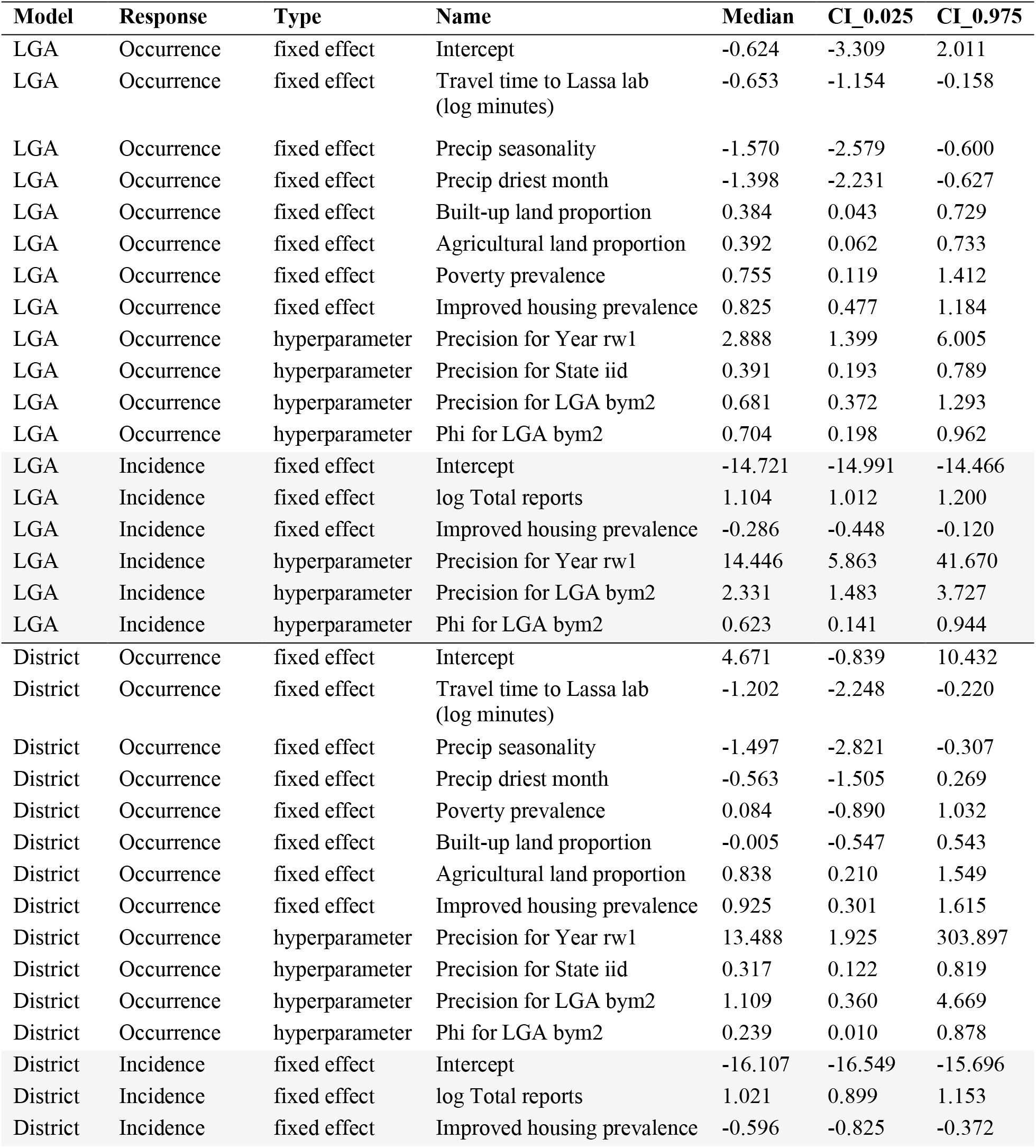
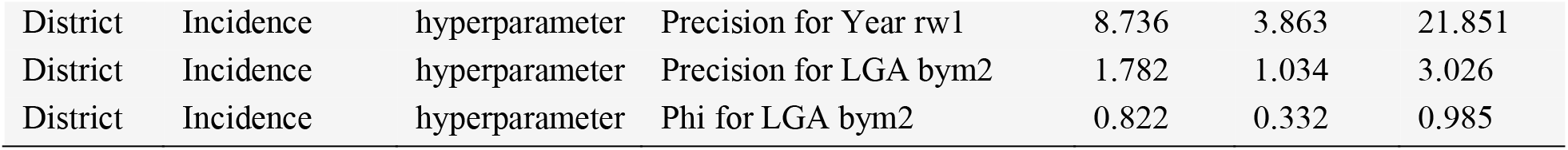
Parameter estimates from the spatial models of Lassa fever occurrence and incidence. Tables show posterior marginal fixed effects parameter and hyperparameter estimates (median and 95% credible interval) from spatial models of annual Lassa fever occurrence and incidence-given-presence at local government authority level (occurrence n=774 LGAs over 4 years; incidence-given-presence n=161 LGAs over 4 years) and at aggregated district level (B) (occurrence n=130 districts over 4 years; incidence-given-presence n=61 districts over 4 years). Occurrence was modelled using a binomial error distribution so estimates are on the log-odds scale, and incidence with Poisson (log link) so estimates are on the natural logarithmic scale. All non-logged covariates were scaled (mean 0, sd 1) prior to model fitting (Methods).

**Supplementary Table 3:** Parameter estimates from the temporal models of Lassa fever incidence in endemic areas. Tables show posterior marginal fixed effects parameter and hyperparameter estimates (median and 95% credible interval) from spatiotemporal models of Lassa fever incidence at district level with climate covariates (South and North). Weekly case counts (n=2090 per model) were modelled using a Poisson likelihood (log-link) so estimates are on the natural logarithmic scale, and all non-logged covariates were scaled (mean 0, sd 1) prior to model fitting (Methods).

| Region | Name | Parameter | Median | CI_0.025 | CI_0.975 |
| --- | --- | --- | --- | --- | --- |
| South | Intercept | fixed effect | -16.458 | -18.3 | -14.589 |
| South | EVI mean daily (120-150 day lag) | fixed effect | 0.73 | 0.529 | 0.932 |
| South | Tmean mean daily (30-60 day lag) | fixed effect | 0.436 | 0.285 | 0.587 |
| South | Precip mean daily (150-180 day lag) | fixed effect | -0.288 | -0.421 | -0.155 |
| South | Precip mean daily (30-60 day lag) | fixed effect | 0.136 | -0.03 | 0.305 |
| South | Precision for District iid random intercept (1/sd) | hyperparam | 430.428 | 10.038 | 375603.7 |
| South | Precision for Year ar1 (1/sd) | hyperparam | 0.165 | 0.078 | 0.33 |
| South | Rho for Year ar1 (correlation coef.) | hyperparam | 0.79 | 0.625 | 0.844 |
| South | Precision for Epi-week ar1 | hyperparam | 1.183 | 0.786 | 1.422 |
| South | Rho for Epi-week ar1 (correlation coef.) | hyperparam | 0.889 | 0.859 | 0.93 |
| North | Intercept | fixed effect | -17.637 | -19.379 | -16.045 |
| North | EVI mean daily (30-60 day lag) | fixed effect | -0.981 | -1.503 | -0.499 |
| North | Precision for District iid random intercept (1/sd) | hyperparam | 10.224 | 0.507 | 744.374 |
| North | Precision for Year ar1 (1/sd) | hyperparam | 0.425 | 0.179 | 0.937 |
| North | Rho for Year ar1 (correlation coef.) | hyperparam | 0.655 | 0.284 | 0.868 |
| North | Precision for Epi-week ar1 | hyperparam | 0.43 | 0.173 | 1.012 |
| North | Rho for Epi-week ar1 (correlation coef.) | hyperparam | 0.948 | 0.873 | 0.98 |

**Supplementary Table 4:** Lassa-fever endemic districts included in the temporal models. The table shows the name and state of each spatially aggregated district included in temporal incidence models, the local government authorities (LGAs) within each district and the total number of confirmed cases detected across all years of surveillance. Cases from 2012-2016 are from the Weekly Epidemiological Reports regime, and from 2017-2019 are from the NCDC Situation Reports surveillance regime.

| District | State | LGAs | Cases total<br>(2012-2019) | Cases 2017-2019 |
| --- | --- | --- | --- | --- |
| South_1 | Edo | Akoko-Edo, Etsako Central, Etsako East, Etsako West, Owan East, Owan West | 215 | 208 |
| South_2 | Edo | Egor, Ikpoba-Okha, Oredo, Ovia North East, Ovia South West | 49 | 42 |
| South_3 | Edo | Esan Central, Esan North East, Esan South East, Esan West, Igueben, Orhionmwon, Uhumwonde | 722 | 340 |
| South_4 | Ondo | Akoko North East, Akoko North West, Akoko South East, Akoko South West, Ose, Owo | 456 | 386 |
| South_5 | Ondo | Akure North, Akure South, Ese-Odo, Idanre, Ifedore, Ilaje, Ile-Oluji-Okeigbo, Irele, Odigbo, Okitipupa, Ondo East, Ondo West | 65 | 51 |
| North_1 | Plateau | Barikin Ladi, Bassa_Plateau, Jos East, Jos North, Jos South, Riyom | 59 | 38 |
| North_2 | Taraba | Ardo-Kola, Jalingo, Karim-Lamido, Lau | 76 | 34 |
| North_3 | Bauchi | Alkaleri, Bauchi, Kirfi | 48 | 33 |
| North_4 | Bauchi | Bogoro, Dass, Tafawa-Balewa | 40 | 16 |
| North_5 | Bauchi | Ningi, Toro | 32 | 30 |

**Supplementary Table 5:** Data sources for all covariates included in analyses. The table includes the sources and rationale (hypothesis) for inclusion of covariates in spatial and spatiotemporal models of Lassa fever incidence across Nigeria. Modelling is described in full in Methods.

| Covariate | Type | Units | Time period | Model | Rationale | Data source, methods | Potential sources of error/misspecification |
| --- | --- | --- | --- | --- | --- | --- | --- |
| Mean distance from laboratory with LF diagnostic capacity (log-transformed) | Con | Km | Annual | Spat., Temp. | Lassa fever detections were historically geographically biased towards areas near to diagnostic laboratories (e.g. ISTH; Gibb *et al* 2017) | Derived from diagnostic laboratory locations (via NCDC) | Distance does not necessarily equate to access and there are different capacities for each laboratory. Health-seeking behaviour will drive small-scale patterns. |
| Mean distance to nearest hospital (log-transformed) | Con | Km | 2015 | Spat. | Proxy for public access to larger/regional healthcare facilities with links to state-level surveillance infrastructure | Estimated as distances from geolocated hospitals in Maina *et al.*, 2019<sup>44</sup> | Distance does not necessarily linearly equate to access. Health-seeking behaviour will drive small-scale patterns. |
| State ID | Cat | n/a | n/a | Spat. | State-level differences in LF surveillance and awareness may influence sensitivity of reporting | n/a | Surveillance and awareness will likely not conform to exact boundaries and spatial scale may be moderately mismatched |
| Air temperature (annual mean and seasonality) | Con | °C | 2011-19, annual | Spat. | Temperature may affect the environmental suitability for *M. natalensis* and/or viral transmission (e.g. through affecting persistence in the environment). | Derived from daily temperature averages from NOAA CPC Precipitation https://www.esrl.noaa.gov/psd/data/gridded/data.cpc.globaltemp.html | Interpolated estimates across state and time will differ slightly from exact values. It is a broad proxy, as food growth, env. stress and viral persistence will all vary with microclimate and host behavioural variation |
| Precipitation (mean wettest month, mean driest month) | Con | mm | 2011-19, annual | Spat. | Evidence of links between *M. natalensis* population ecology and rainfall seasonality, and past evidence that human LF is linked to areas of high rainfall<sup>12</sup> | Derived from daily Africa precipitation rasters from CHIRPS (Climate Hazards Infrared Precipitation with Stations)<sup>41</sup> | Interpolated estimates across state and time will differ slightly from exact values. It is a broad proxy, as food growth, env. stress and viral persistence will all vary with microclimate and host behavioural variation |
| Precipitation annual seasonality (coefficient of variation) | Con | mm | 2011-19, monthly | Spat. | Evidence of links between *M. natalensis* population ecology and rainfall seasonality, and past evidence that human LF is linked to areas of high rainfall<sup>12</sup> | Derived from daily Africa precipitation rasters from CHIRPS (Climate Hazards Infrared Precipitation with Stations)<sup>41</sup> | Interpolated estimates across state and time will differ slightly from exact values. It is a broad proxy, as food growth, env. stress and viral persistence will all vary with microclimate and host behavioural variation |
| Mean precipitation in 60-day window (starting 0, 30, 60, 90, 120 day lag prior to reporting week) | Con | mm | 2011-20, weekly | Temp. | Evidence that *M. natalensis* population densities and patterns of human exposure may be linked to rainfall patterns<sup>12</sup> | Derived from smoothed daily Africa precipitation rasters from CHIRPS<sup>41</sup> | Interpolated estimates across state and time will differ slightly from exact values. It is a broad proxy, as food growth, env. stress and viral persistence will all vary with microclimate and host behavioural variation |
| Mean, min and max air temperature in 60-day window (starting 0, 30, 60, 90, 120 day lag prior to reporting week) | Con | °C | 2011-20, weekly | Temp. | *M. natalensis* population densities, viral persistence and patterns of human exposure may be linked to climatic patterns<sup>12</sup> | Derived from smoothed daily Tmin and Tmax temperature layers from NOAA CPC | Interpolated estimates across state and time will differ slightly from exact values. It is a broad proxy, as food growth, env. stress and viral persistence will all vary with microclimate and host behavioural variation |
| Mean EVI in 60-day window (starting 0, 30, 60, 90, 120 day lag prior to reporting week) | Con | EVI | 2011-20, weekly | Temp. | Evidence that *M. natalensis* population densities and patterns of human exposure may be linked to climatically-driven vegetation dynamics (i.e. due to seasonal resource availability)<sup>12</sup> | Derived from processed and smoothed 16-day EVI layers from NASA | Interpolated estimates across state and time will differ slightly from exact values. It is a broad proxy, as food growth, env. stress and viral persistence will all vary with microclimate and host behavioural variation |
| Urban land cover | Con | Prop. area | 2015 | Spat. | May impact both *M. natalensis* densities (which are often lower in highly urbanised environments) and human access to healthcare facilities (i.e. detection probability) | ESA-CCI Land Cover 2015 300m raster (data for a single year as relatively low change in proportion cover during surveillance period) https://www.esa-landcover-cci.org | Gross land cover categorisation will not fully capture the high variation and difference between difference habitat types. The quality of the habitats, their fragmentation and the geographical differences in such habitats will also likely be important. |
| Agricultural land cover | Con | Prop. area | 2015 | Spat. | *M. natalensis* densities often observed to be highest in agricultural settings<sup>29,49</sup> and cropping and crop preparation practices are a hypothesised driver of human-rodent contact<sup>12</sup> | ESA-CCI Land Cover 2015 (data for a single year as relatively low change in proportion cover during surveillance period) https://www.esa-landcover-cci.org/ | Gross land cover categorisation will not fully capture the high variation and difference between difference habitat types. The quality of the habitats, their fragmentation and the geographical differences in such habitats will also likely be important. |
| Total human population (log transformed) | Con | No. people | 2012-20, annual | Spat., Temp. | Controlling for effects of increased human population on potential for LASV exposure. | WorldPop | Does not account for exact proportion of people that at-risk and how this varies over space and time. More accurate might be density of agricultural workers for instance. |
| Proportion of LGA population living in poverty (<\$1.25 per day) | Con | Prop. area | 2010 | Spat. | Evidence that human LASV exposure may often be linked to ability to store food in rodent-proof containers and ability to rodent-proof housing<sup>8,53</sup> | WorldPop | Poverty is a broad proxy for ability for people to prevent or react to disease risk. Differences e.g. in awareness, personal choice and beliefs, and personal circumstances will influence actions. |
| Proportion of population in locales with improved housing | Con | Prop. area | 2015 | Spat. | Evidence that human LASV exposure may be linked to ability to rodent-proof housing<sup>53</sup> | Derived from Malaria Atlas Project modelled data on prevalence of improved housing<sup>42</sup> (5km<sup>2</sup> resolution) and human population layers (WorldPop) | This index intends to capture the ability of rodents to exploit food in houses but the actual design and use of house will have strong impact on how many host individuals are able to invade. |

## References

1. Siddle, K. J. et al. Genomic Analysis of Lassa Virus during an Increase in Cases in Nigeria in 2018. N. Engl. J. Med. NEJMoa1804498 (2018) doi:10.1056/NEJMoa1804498.

2. Kafetzopoulou, L. E. et al. Metagenomic sequencing at the epicenter of the Nigeria 2018 Lassa fever outbreak. Science 80, 74–77 (2019).

3. Gibb, R., Moses, L. M., Redding, D. W. & Jones, K. E. Understanding the cryptic nature of Lassa fever in West Africa. Pathog. Glob. Health 111, 276–288 (2017).

4. Nigeria Centre for Disease Control, Lassa fever Situation Report, 12 April 2020. https://ncdc.gov.ng/themes/common/files/sitreps/8c02d1bf6e3e02aa2adfe144dda40db2.pdf (x2020).

5. Ipadeola, O. et al. Epidemiology and case-control study of Lassa fever outbreak in Nigeria from 2018 to 2019. J. Infect. 80, 578–606 (2020).

6. Rottingen, J. et al. New vaccines against epidemic infectious diseases. New Engl. J. Med. 376, 610–613 (2017).

7. Lo Iacono, G. et al. Using modelling to disentangle the relative contributions of zoonotic and anthroponotic transmission: the case of Lassa fever. PLoS Negl. Trop. Dis. 9, e3398 (2015).

8. Andersen, K. G. et al. Clinical sequencing uncovers origins and evolution of Lassa Virus. Cell 162, 738–750 (2015).

9. Ilori, E. A. et al. Epidemiologic and Clinical Features of Lassa Fever Outbreak in Nigeria, January 1-May 6, 2018. Emerg. Infect. Dis. 25, 1066–1074 (2019).

10. Bonwitt, J. et al. At Home with Mastomys and Rattus: Human–Rodent Interactions and Potential for Primary Transmission of Lassa Virus in Domestic Spaces. Am. J. Trop. Med. Hyg. 96, 935–943 (2017).

11. Dzingirai, V. et al. Structural drivers of vulnerability to zoonotic disease in Africa. Philos. Trans. R. Soc. B 372, 20160169 (2017).

12. Fichet-Calvet, E. & Rogers, D. J. Risk maps of Lassa fever in West Africa. PLoS Negl. Trop. Dis. 3, e388 (2009).

13. Redding, D. W., Moses, L. M., Cunningham, A. A., Wood, J. & Jones, K. E. Environmental-mechanistic modelling of the impact of global change on human zoonotic disease emergence: a case study of Lassa fever. Methods Ecol. Evol. 7, 646–655 (2016).

14. McCormick, J. B., Webb, P. A., Krebs, J. W., Johnson, K. M. & Smith, E. S. A prospective study of the epidemiology and ecology of Lassa fever. J. Infect. Dis. 155, 437–444 (1987).

15. Bausch, D. G. et al. Lassa Fever in Guinea: I. Epidemiology of human disease and clinical observations. Vector Borne Zoonotic Dis. 1, 269–281 (2001).

16. Wilkinson, A. Beyond biosecurity: the politics of Lassa fever in Sierra Leone. in One Health: Science, Politics and Zoonotic Disease in Africa (ed. Bardosh, K.) 117–138 (Routledge: London, NY, 2016).

17. Sogoba, N. et al. Lassa Virus seroprevalence in Sibirilia Commune, Bougouni District, Southern Mali. Emerg. Infect. Dis. 22, 657–663 (2016).

18. Akpede, G. O., Asogun, D. A., Okogbenin, S. A. & Okokhere, P. O. Lassa fever outbreaks in Nigeria. Expert Rev. Anti. Infect. Ther. 16, 663–666 (2018).

19. Asogun, D. A. et al. Molecular Diagnostics for Lassa Fever at Irrua Specialist Teaching Hospital, Nigeria: Lessons Learnt from Two Years of Laboratory Operation. PLoS Negl. Trop. Dis. 6, (2012).

20. Akpede, G. O. et al. Caseload and Case Fatality of Lassa Fever in Nigeria, 2001– 2018: A Specialist Center’s Experience and Its Implications. Front. Public Heal. 7, 170 (2019).

21. Shaffer, J. G. et al. Lassa fever in post-conflict Sierra Leone. PLoS Negl. Trop. Dis. 8, e2748 (2014).

22. Ilori, E. A. et al. Increase in Lassa Fever Cases in Nigeria, January–March 2018. Emerg. Infect. Dis. 24, 2018–2019 (2019).

23. NCDC. First Annual Report of the Nigeria Centre for Disease Control. http://www.ncdc.gov.ng/themes/common/docs/protocols/78_1515412191.pdf (2016).

24. Fichet-Calvet, E., Becker-Ziaja, B., Koivogui, L. & Günther, S. Lassa serology in natural populations of rodents and horizontal transmission. Vector-Borne Zoonotic Dis. 14, 665–74 (2014).

25. Fichet-Calvet, E., Lecompte, E., Koivogui, L., Daffis, S. & Meulen, J. T. Reproductive characteristics of Mastomys natalensis and Lassa virus prevalence in Guinea, West Africa. Vector-Borne Zoonotic Dis. 8, 41–48 (2008).

26. Olayemi, A. et al. Arenavirus Diversity and Phylogeography of Mastomys natalensisRodents, Nigeria. Emerg. Infect. Dis. 22, 687–690 (2016).

27. Olayemi, A. et al. Small mammal diversity and dynamics within Nigeria, with emphasis on reservoirs of the lassa virus. Syst. Biodivers. 16, 118–127 (2018).

28. Ehichioya, D. U. et al. Phylogeography of Lassa Virus in Nigeria. J. Virol. 93, e00929–19 (2019).

29. Leirs, H., Verhagen, R., Verheyen, W., Mwanjabe, P. & Mbise, T. Forecasting Rodent Outbreaks in Africa: An Ecological Basis for Mastomys Control in Tanzania. J. Appl. Ecol. 33, 937–943 (1996).

30. Massawe, A. W., Rwamugira, W., Leirs, H., Makundi, R. H. & Mulungu, L. S. Do farming practices influence population dynamics of rodents? A case study of the multimammate field rats, Mastomys natalensis, in Tanzania. Afr. J. Ecol. 45, 293–301 (2007).

31. Ballester, J., Lowe, R., Diggle, P. J. & Rodó, X. Seasonal forecasting and health impact models: Challenges and opportunities. Ann. N. Y. Acad. Sci. 1382, 8–20 (2016).

32. Lowe, R. et al. Evaluating probabilistic dengue risk forecasts from a prototype early warning system for Brazil. Elife 5, 1–18 (2016).

## Methods References

33. Dan-Nwafor, C. C. et al. Measures to control protracted large Lassa fever outbreak in Nigeria, 1 January to 28 April 2019. Eurosurveillance 24, 1–4 (2019).

34. Zhao, S. et al. Large-scale Lassa fever outbreaks in Nigeria: quantifying the association between disease reproduction number and local rainfall. bioRxiv Prepr. http://dx.doi.org/10.1101/60270 1–13 (2019).

35. Akhmetzhanov, A. R., Asai, Y. & Nishiura, H. Quantifying the seasonal drivers of transmission for Lassa fever in Nigeria. Phil. Trans. R. Soc. B 374, 20180268 (2019).

36. Gotway, C. A. & Young, L. J. Combining incompatible spatial data. J. Am. Stat. Assoc. 97, 632–648 (2002).

37. Lindgren, F. & Rue, H. Bayesian Statistical Modelling with R-INLA. J. Stat. Softw. 63, (2010).

38. Hijmans, R. J. raster: Geographic data analysis and modelling. R package v.2.5-8. https://CRAN.R-project.org/package=raster. (2016).

39. Hunziker, P. velox: Fast Raster Manipulation and Extraction. R package v.0.2.0. https://CRAN.R-project.org/package=velox. (2017).

40. Rue, H., Martino, S. & Chopin, N. Approximate Bayesian inference for latent Gaussian models by using integrated nested Laplace approximations. J. R. Stat. Soc. Ser. B Stat. Methodol. 71, 319–392 (2009).

41. Redding, D. W., Lucas, T. C. D., Blackburn, T. M. & Jones, K. E. Evaluating Bayesian spatial methods for modelling species distributions with clumped and restricted occurrence data. PLoS One 12, e0187602 (2017).

42. Maina, J. et al. A spatial database of health facilities managed by the public health sector in sub Saharan Africa. Sci. data 6, 134 (2019).

43. Funk, C. et al. The climate hazards infrared precipitation with stations - A new environmental record for monitoring extremes. Sci. Data 2, 1–21 (2015).

44. Watanabe, S. Asymptotic Equivalence of Bayes Cross Validation and Widely Applicable Information Criterion in Singular Learning Theory. J. Mach. Learn. Res. 11, 3571–3594 (2010).

45. Gelman, A., Hwang, J. & Vehtari, A. Understanding predictive information criteria for Bayesian models. Stat. Comput. 24, 997–1016 (2014).

46. Hooten, M. B. & Hobbs, N. T. A guide to Bayesian model selection for ecologists. Ecol. Monogr. 85, 3–28 (2015).

47. Kelly, A. H. & Marí Sáez, A. Shadowlands and dark corners: An anthropology of light and zoonosis. Med. Anthropol. Theory 5, 21–47 (2018).

48. Fichet-Calvet, E. et al. Fluctuation of abundance and Lassa virus prevalence in Mastomys natalensis in Guinea, West Africa. Vector-Borne Zoonotic Dis. 7, 119–28 (2007).

49. Makundi, R. H., Massawe, A. W. & Mulungu, L. S. Reproduction and population dynamics of Mastomys natalensis Smith, 1834 in an agricultural landscape in the Western Usambara Mountains, Tanzania. Integr. Zool. 2, 233–238 (2007).

50. FEWSNET. Famine Early Warning Systems Network: Revised livelihoods zone map and descriptions for Nigeria. https://fews.net/west-africa/nigeria/livelihood-description/september-2018 (2018).

51. Lowe, R. et al. Nonlinear and delayed impacts of climate on dengue risk in Barbados: A modelling study. PLoS Med. 15, 1–24 (2018).

52. Mari Saez, A. et al. Rodent control to fight Lassa fever: Evaluation and lessons learned from a 4-year study in Upper Guinea. PLoS Negl. Trop. Dis. 12, 1–16 (2018).

53. Tusting, L. S. et al. Mapping changes in housing in sub-Saharan Africa from 2000 to 2015. Nature (2019) doi:10.1038/s41586-019-1050-5.

